# Recessive genomic and phenotypic variation in consanguineous families with cerebral palsy

**DOI:** 10.1101/2025.11.04.25339178

**Authors:** Pritha Bisarad, Yung-Chun Wang, Peter T. Skidmore, Carolina I. Galaz-Montoya, Sara A. Lewis, Bader Alhaddad, Nahyun Kong, Dominic Julian, Helen Magee, Tyler N. Kruer, Yuhan Xie, Wangjie Zheng, Boyang Li, Fatemeh V. Rajabpour, James Liu, Anjali Revanur, Khadijah Bakur, Saghar Ghasemi Firouzabadi, Sarina Sharbatkhori, Abbas Tafakhori, Ehsan Taghiabadi, Ermia Nezaminargabad, Shohreh Vosoogh, Javad Jamshidi, Serajaddin Arefnia, Seyed Ahmad Hosseini, Alireza Khajehmirzaei, Faezeh Jamali, Azadeh Ahmadifard, Hamidreza Khodadadi, Parvaneh Daneshmand, Saeed Bohlega, Sateesh Maddirevula, Seba Saleh Nadeef, Mais O. Hashem, Mustafa A. Salih, Inaam N. Mohmed, Heinrich Sticht, Sara Peres Morias, Joana Damásio, Mariana Santos, José Leal Loureiro, Rita Rodrigues, Giovanni Stevanin, Mehdi Benkirane, Benjamin Dauriat, Nicholas Head, Júlia Baptista, Saeid Shahhosseini, Farhan Mohammad, Hongyu Zhao, Sergio Padilla-Lopez, Fowzan Alkuraya, Somayeh Bakhtiari, Michael C. Kruer, Sheng Chih Jin, Hossein Darvish

## Abstract

Cerebral palsy (CP) is a neurodevelopmental disorder of motor function, with genetic etiologies, particularly *de novo* variants, identified in approximately one-third of cases. The contribution of consanguinity – long-recognized as a CP risk factor – has remained undefined. Here, we report findings from 188 primarily consanguineous Middle Eastern families with CP and identified putative causative genes in nearly three-quarters. The majority demonstrated recessive inheritance, although multi-level consanguinity and multilocus pathogenic variants complicated Mendelian assortment analyses. We identified 110 known CP-associated genes – five with phenotypic expansions and three others exhibiting new recessive inheritance patterns – and 24 novel candidates. We characterized ten candidates as high-confidence based on independent replication and protein modeling. We experimentally validated a role for *SUCO* variants in CP and newly identified a role for mid-gestational migrating excitatory neurons in the disorder. These findings highlight new genes, pathways, and phenotypes that reveal striking genomic diversity in CP.

## INTRODUCTION

Cerebral palsy (CP) is a major neurodevelopmental disorder impacting motor function, affecting an estimated one in three hundred children.^1^ It is the most common cause of physical disability in childhood, and it is a lifelong condition.^2,3^ As a developmental disorder of movement, individuals with CP may experience difficulties with walking, speech, feeding, and fine motor skills.

Although CP has historically been associated with prenatal and perinatal injury^4^, individually rare monogenic and copy number variants have been implicated in a substantial proportion of individuals with CP.^5–8^ Meta-analyses report etiologic yields of whole exome or whole genome sequencing in approximately one-third of cases, comparable to the etiologic yields of genome sequencing for other neurodevelopmental disorders such as autism and intellectual disability/global developmental delay.^9–11^ Despite these advances, for most individuals with CP for whom a genetic diagnosis is suspected^12^, no etiology is able to be detected. In addition, a disease association has been established for ∼30% of human genes by the Online Mendelian Inheritance in Man database. These observations suggest that many additional CP-associated genes await discovery.

Prior genomic analyses in CP, largely conducted in Chinese or Western cohorts, identified a major role for *de novo* variants (DNVs) in CP, potentially accounting for the predominantly sporadic occurrence of the disorder.^8^ Approximately one-third of CP-associated genes are inherited in a recessive pattern in Western populations, and this proportion may be even higher in consanguineous populations.^13^ Globally, an estimated 10% of individuals are born to consanguineous unions^14^, with regional rates as high as 40% in Saudi Arabia^15^, with similar frequencies in Iran^16^, Egypt^17^, slightly higher rates in the United Arab Emirates^18^, and slightly lower rates in Turkey^19^. However, while consanguinity has long been recognized as a CP risk factor^20,21^, its contribution to the genomic architecture of CP has been poorly defined. Studies of consanguineous populations in other neurodevelopmental disorders (NDDs) have helped characterize the full spectrum of human genomic diversity, uncovering new disease associated genes and novel and expanded phenotypes. Our prior work in congenital heart disease indicated that distinct biological mechanisms distinguish recessively-acting variants (ciliary pathways) from those exhibiting *de novo* patterns (chromatin modifiers acting via haploinsufficiency).^22^ This suggests disease gene discovery focused on recessive genotypes might not only uncover additional genes and phenotypes but also help identify important biological mechanisms for CP that complement those implicated by other forms of inheritance.

Here, we present results from a large cohort analysis of CP in families of Middle Eastern origin. Our analyses reveal substantial genetic and phenotypic heterogeneity and highlight the value of studying populations of different ancestry for a more complete understanding of genomic contributions to CP.

## RESULTS

### The multiplex, autozygous populations in cerebral palsy (MAP CP) cohort

We enrolled 345 individuals with CP from 188 families of Middle Eastern origin (**Supplementary Table 1**). The majority of families (86.17%; 162/188) reported consanguinity, and 76.06% of families (143/188) were multiplex, with two or more affected children with CP (**Figure 1a**). Among affected individuals, the primary motor disorder was most commonly spasticity, followed by hypotonia, dyskinesia – dystonia, ataxia, and dyskinesia – chorea/ballism (**Figure 1b**), similar to published data for CP at large.^23^ These disorders most often manifested in a generalized or quadriplegic distribution (**Figure 1c**); cases in our cohort were more likely to be quadriplegic/generalized than observed in registry-based^24^ surveys (χ² = 103.24, p<0.0001), suggesting more severe involvement. Brain MRI findings were classified using the MRI Classification System (MRICS; **Figure 1d**). Comorbidities, including intellectual disability/global developmental delay, epilepsy, autism, and hydrocephalus were frequently seen and reported in **Supplementary Table 1**.

**Figure 1.**
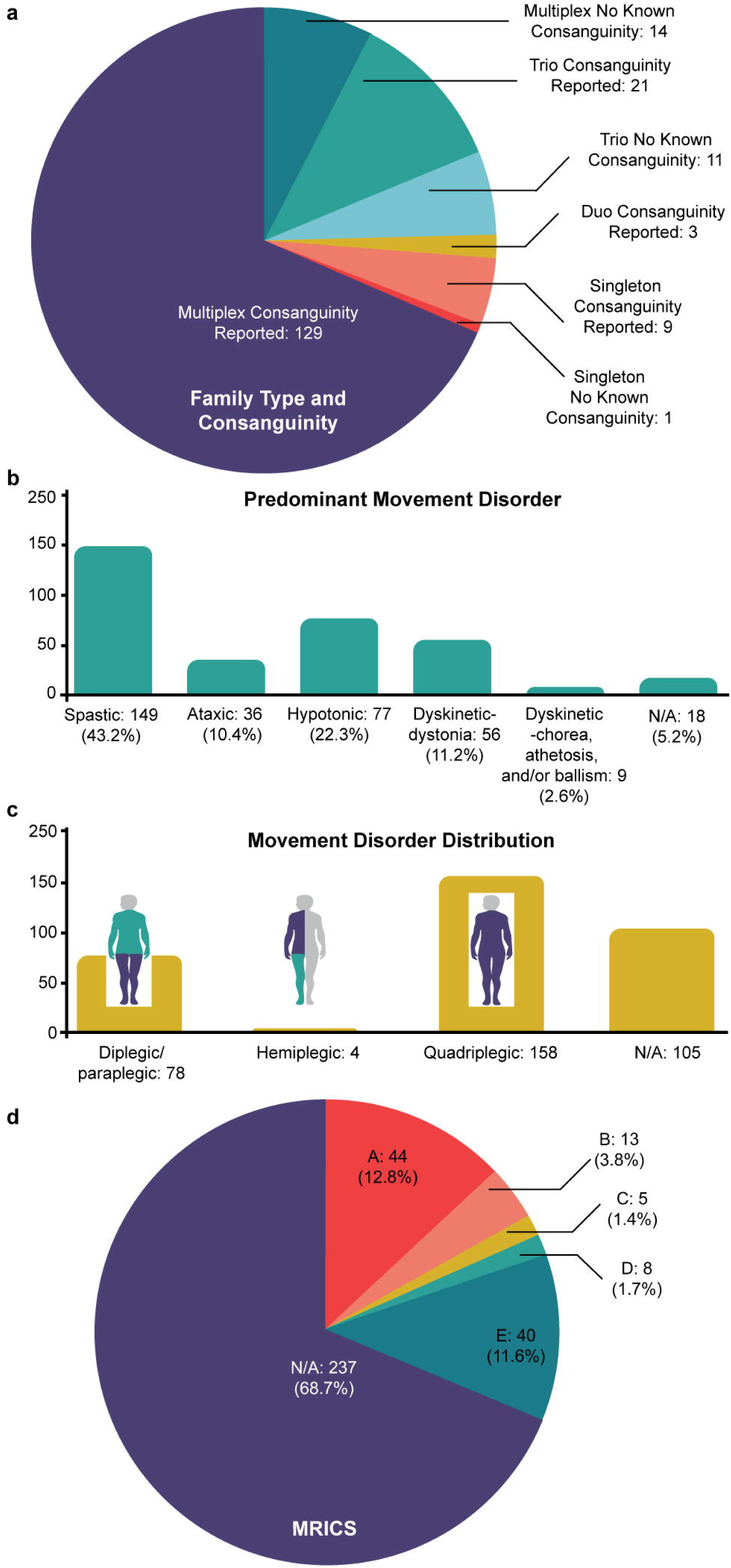
Summary of Cohort. **a.** Proportion of reported consanguinity superimposed with proportion of multiplex families (n = 188 families). **b.** Predominant movement disorder by proportion (n = 345 probands). **c.** Movement disorder distribution by shaded body region (n = 345 probands). **d.** Brain MRI findings summarized by MRI Classification System (MRICS).

### Variant analysis

We performed whole-exome sequencing in all families and called single nucleotide variants (SNVs), small insertions/deletions (INDELs) and copy number variants as previously described.^8^ Segregation analysis was used to prioritize X-linked, autosomal recessive, and DNVs. Sequencing quality metrics can be found in **Supplementary Table 2**. Variant calling thresholds and filtering criteria are detailed in the **Methods**. Most public reference databases, including gnomAD^25,26^, ExAC^27,28^, and TOPMed^29^ are predominantly composed of individuals with European ancestry, so we incorporated the Greater Middle Eastern (GME) Variome database^30^ to better evaluate population-specific allele frequencies (**Supplementary Table 3**).

### Autozygosity mapping & candidate gene prioritization

Identity (IBD) by descent occurs when a phased haplotype block is inherited from an ancestor without recombination. In consanguineous families, IBD haplotypes segregate through both sides of a family due to a common ancestor. When these IBD haplotypes co-occur in a descendent, this leads to autozygosity characterized by runs of homozygosity (ROH). Historically, autozygosity mapping has shown utility in recessive disease gene discovery since ROH regions are enriched for deleterious homozygous alleles inherited from the same shared relative^31,32^ but autozygosity mapping has not previously been applied to CP.

Worldwide and in our cohort, first cousin marriages are the most common form of consanguineous union.^33^ Prior work has shown that incorporating autozygosity mapping into variant interpretation workflows improves the predictive power to identify a causative gene, with larger ROH regions most likely to reflect recent consanguinity and harbor disease-associated genes.^34^ Smaller regions of ROH (i.e. <1 Mb) typically reflect ancestral regions that have been subjected to selective pressures for longer periods of time and are thus less likely to harbor disease-associated variants.^35^ However, recent studies have also reported that causative variants often reside outside of the largest ROH blocks in consanguineous populations for unclear reasons.^36^

Given the nature of our cohort, we incorporated autozygosity mapping into our variant calling and interpretation pipeline, using a custom algorithm to visualize variants located within ROH regions ≥ 1 Mb. We compared total ROH burden across our cohort (**Supplementary Table 4**). We observed greater ROH burden in families reporting consanguinity versus those denying it (*p* = 2.1×10^-9^), but also noted a higher burden of ROH in MAP CP families denying consanguinity compared to those in a US-based cohort (*p* < 3.9×10^-3^; **Figure 2a**), indicating that consanguineous relationships may exist that families are unaware of. We then compared total ROH burden against reported consanguinity. We found that observed values exceeded pedigree-derived expected values (*p* < 6.8 x10^-6^), and for 77.3% of families included in this analysis, ROH burden was ≥ 2 standard deviations above expectation, indicating that degrees of consanguinity unrecognized by families were present (**Figure 2b**). While most putative causative variants (known or novel gene prioritized in each family) were found within the largest ROH blocks as predicted based on degree of consanguinity, a notable proportion (46.96%) mapped to smaller but recent ROH segments (**Figure 2c**). Furthermore, the integration of segregation data from multiple affected individuals with ROH analyses revealed that 68.69% of causative variants were found in ROH regions conserved across affected siblings (**Figure 2d**), similar to that observed in other multiplex, consanguineous cohorts.^29^

**Figure 2.**
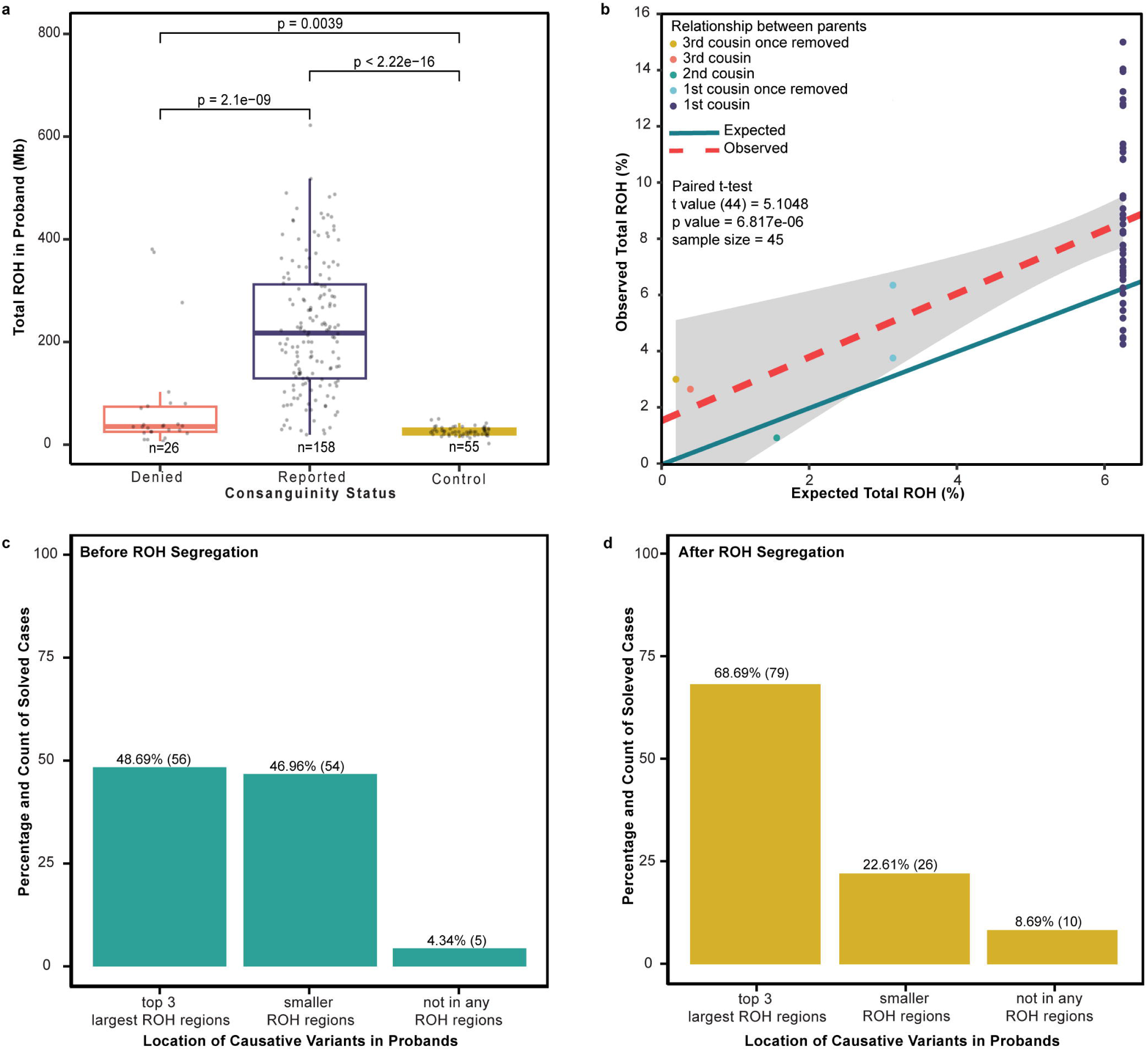
Autozygosity mapping and candidate gene prioritization. **a.** Comparison of total calculated ROH regions in families that denied consanguinity (n=26), families that reported consanguinity (n=158), and a non-consanguineous control cohort from the United States (n=55). **b.** Calculated vs. expected total ROH burdens in families based on available pedigree information (n=45) shows higher ROH burden than expected (significance assessed by paired t-test). **c.** Percent distribution of causative homozygous recessive (missense) variants based on the size of the ROH region harboring that variant in the proband. **d.** Percent distribution of causative homozygous recessive (missense) variants based on the size of the ROH region harboring that variant in the proband after segregation.

### Identification of CP-associated genes

Variant classification was performed by multidisciplinary review, incorporating inheritance patterns, predicted variant effects, and phenotypes using American College of Medical Genetics and Genomics (ACMG) criteria (see **Methods**).^37^ We classified genes as putative causative known disease-associated genes based on the presence of Likely Pathogenic or Pathogenic variants with appropriate inheritance patterns and a concordant phenotype based on established Online Mendelian Inheritance in Man (OMIM)^38^ disease associations. We identified 102 putative causative known genes: 91 autosomal recessive (86 homozygous, 5 compound heterozygous), 8 *de novo*, and 3 demonstrating X-linked recessive inheritance. We identified 5 additional genes that represented known disease-associated genes with phenotypic expansions and 4 more known disease-associated genes that demonstrated a recessive inheritance pattern distinct from the typical dominant pattern of inheritance. We identified 24 novel disease-associated candidate genes (**Supplementary Table 5**); 20 were homozygous, 3 were compound heterozygous, and one demonstrated X-linked recessive inheritance. We identified 9 genes as high-confidence candidate genes based on *in silico* structural modeling and/or independent replication (*SYNRG*, *SFPQ*) and one additional gene (*SUCO*) as an experimentally validated disease-associated gene (see below). In total, we identified 120 CP-associated genes (**Supplementary Table 6**) – 110 with known disease associations with some demonstrating novel phenotypes or inheritance patterns, 9 high-confidence candidates, and one newly-validated disease gene.

### Etiologic yield in participants

Putative causative genes (**Supplementary Table 3**) reflecting likely monogenic etiologies (a CP-associated gene or candidate gene demonstrating appropriate segregation) were identified in 137 families in our cohort (137/188; 72.87%) across CP subtypes (**Figure 3a**). This yield is significantly higher than the yield reported for exome sequencing from recent meta-analyses of CP cases^9,13^, likely reflecting the consanguineous family structure and high proportion of multiplex families in our cohort. For 83.94% (115/137) of families with putative causative genes, variants were homozygous recessive, while for 7.29% (10/137) variants were compound heterozygous. Just under 3% of families (4/137) exhibited X-linked hemizygous inheritance, and 5.83% (8/137) of families had putative causative DNVs. Among these 137 families for whom a likely monogenic cause was identified, the same causative gene was detected in two or more families 16 times. Novel CP candidate genes segregated in 24 families with one gene, *ARHGEF11*, recurrently detected in two families. In contrast to our prior findings in a predominantly Western cohort, in this Middle Eastern cohort, we did not observe a significant enrichment of DNVs across functional classes (**Supplementary Table 7, Supplementary Figure 1**).

**Figure 3.**
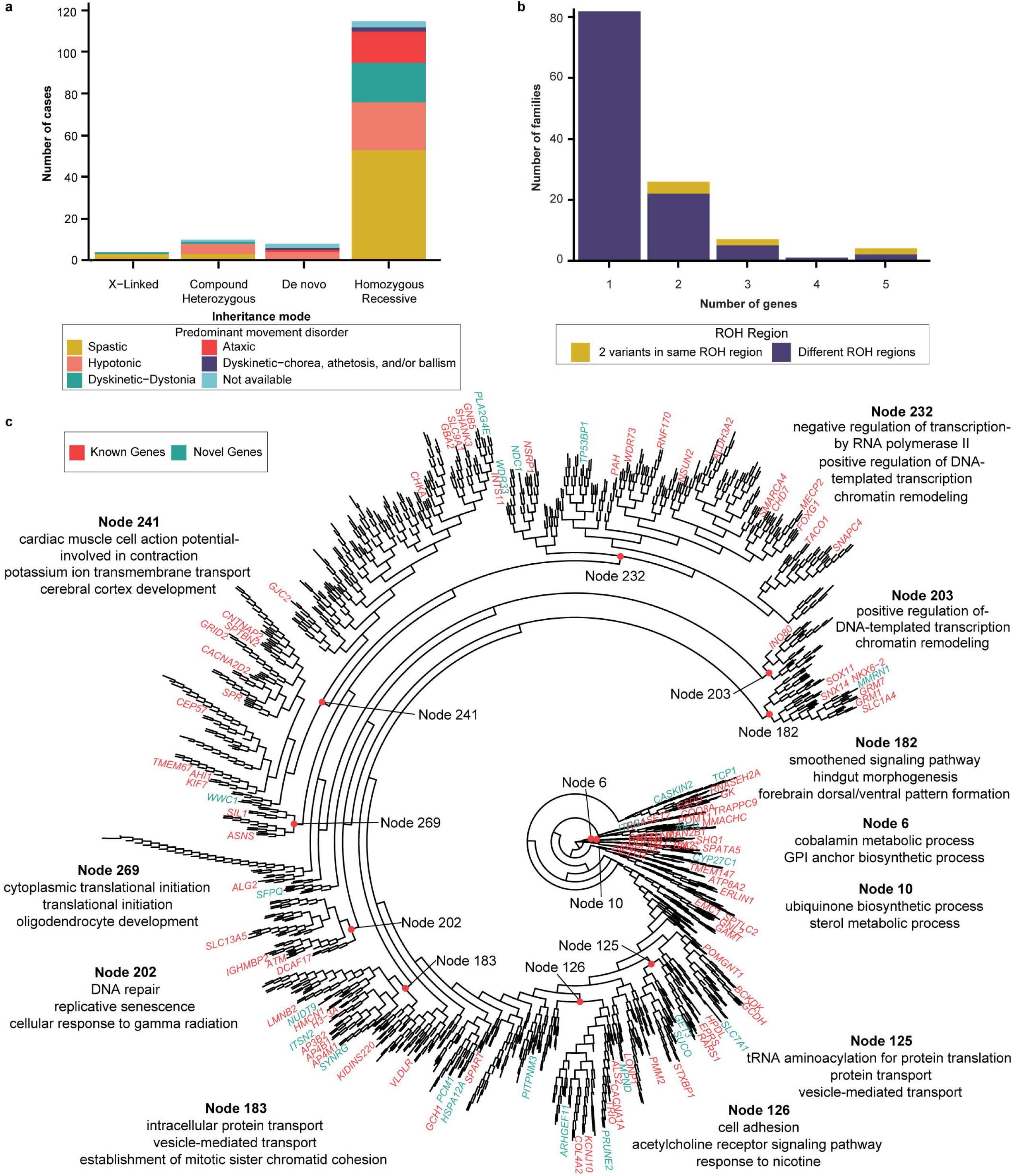
Genes identified in cohort. **a.** Known disease-associated genes stratified by inheritance mode and predominant movement disorder of the proband. **b.** Potential multilocus pathogenic variants were identified in families appropriately segregating two or more genes with damaging variants in known disease-associated genes with compatible phenotypes. **c.** Functional genomic alignment of human gene connectome (HGC) distances between known CP-associated genes and genes identified in the current cohort. HGC distances generated for CP-associated genes (black), known disease associated genes identified in study (red), and CP candidate genes identified in study (green). GO terms for selected nodes obtained from DAVID and summarized using REVIGO shown.

### Multilocus pathogenic variants (MPVs)

In Western populations, for most individuals with a Mendelian disorder, a single causative gene is found; yet in ∼5% of cases, two or more genes with Likely Pathogenic or Pathogenic variants contribute to their phenotype.^39^ In consanguineous populations, recessive genotypes can manifest as homozygous multilocus pathogenic variants (MPVs)^40^, leading to overlapping or blended phenotypes. In our cohort, potential MPVs were detected in 27.8% (38/137) of the families with at least one likely monogenic etiology (**Figure 3b**), similar to rates previously reported in Turkish and Saudi cohorts.^40,41^ Additionally, we observed some of these MPVs segregating in the same ROH region, consistent with autozygosity-derived linkage disequilibrium.

### Human genomic connectome of CP

We performed a functional genomic alignment analysis based on STRING protein-protein interaction relationships to visualize relationships between both known disease-associated and novel candidate genes identified in our cohort and genes previously reported in association with CP. We found that both the known and novel genes we identified were well-represented within enriched CP gene networks (**Figure 3c**). Functional annotation identified important roles for intracellular protein transport [node 183]), chromatin remodeling [nodes 203, 232], transcriptional regulation [nodes 203, 232], tRNA aminoacylation [node 125], protein translation [node 269], sterol metabolism [node 10], and glycosylphosphatidylinositol (GPI) anchor biosynthesis [node 6] within enriched gene clusters. These findings demonstrate converging functional connections between genes across inheritance patterns identified in our Middle Eastern cohort and prior cohorts from the United States, Australia, and China.

### *In silico* prioritization of novel CP-associated candidate genes

We identified 24 novel disease candidate genes (**Supplementary Table 5**) in our cohort and applied a suite of complementary structural variant analyses to verify candidate genes and known CP-associated genes demonstrating alternative inheritance patterns. We incorporated analyses of ΔΔG, deep learning, normal mode analysis, graph signature modeling, statistical potentials, forward/reverse pairing, structural rules, and protein-protein interaction sites/binding potential in a multimodal manner (**Figure 4a, Supplementary Figures 2 & 3**). We assessed 18 genes with homozygous missense variants using this approach and designated 3 genes (*CASKIN2*, *SLC7A1*, and *GET3*) as high-confidence candidate genes based on consistent results across tools (**Supplementary Table 8**).

**Figure 4.**
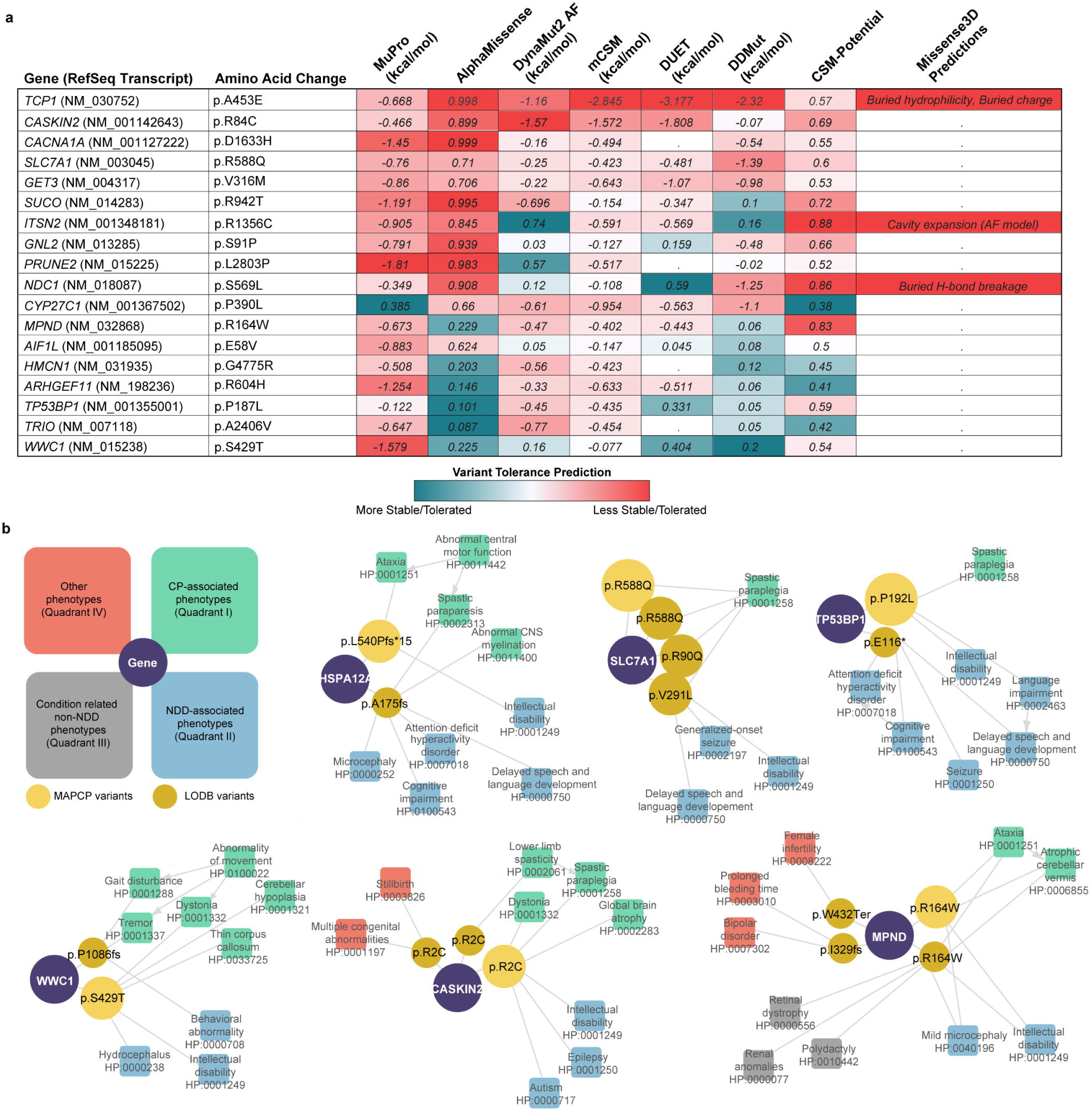
Predicting pathogenicity of high confidence candidate genes using protein models. **a.** Predictions of stability and tolerance of amino acid substitutions in proteins using protein sequence (MuPro) and computed protein models (AlphaMissense, DynaMut2, mCSM, DUET, DDMut, CSM-Potential, Missense3D) for 18 novel and new inheritance genes. Red highlights that substitutions are less tolerated/stable. Blue highlights that substitutions are more tolerated/stable. “.” indicates unavailable data. **b.** Network graphs showing phenotypic distribution and convergence between cases with MAP CP variants (light yellow) and LODB^42^ variants (dark yellow) across six novel genes (purple). Schematic diagram on top left represents broad phenotype classification for each phenotype present. Variant node circle size corresponds to number of individuals with the particular variant.

### Independent replication of novel genes by leveraging a large scale Saudi Arabian dataset

We also evaluated our novel genes by comparing them to hits from the recently developed Saudi Arabian large-scale combined clinical exome/genome sequencing data (Lifera Omics Databank, LODB).^42^ Inheritance, allele frequencies, deleteriousness, and phenotypic convergence were assessed for the 24 novel genes from our cohort in the LODB. This comparative analysis led to additional evidence in support of six candidate genes from our cohort (**Figure 4b, Supplementary Table 9**). Recessively inherited cases with consistent neurodevelopmental and movement disorder phenotypes similar to CP were identified for four genes (*HSPA12A, SLC7A1, TP53BP1, WWC1).* Phenotypically similar cases with homozygous recessive variants were also identified in *CASKIN2* and *MPND*, although pleiotropic phenotypes were also observed (**Supplementary Table 9**). Independent observations in the LODB with convergent phenotypes thus supported four additional high confidence candidate genes (*HSPA12A, MPND, TP53BP1, WWC1)* not already prioritized by protein modeling.

### Experimental validation supports a role for damaging recessive *TCP1* variants in CP

*TCP1* (*CCT1*) encodes a subunit of the eukaryotic TRiC chaperonin complex. TriC is a double ring “folding barrel” with 8 different subunits (CCT1-CCT8) per ring. TriC clients include several proteins crucial for neurodevelopment, including actins and tubulins (previously associated with genetic forms of CP^8^), heterotrimeric G-protein β subunits (*GNB1* is a CP-associated gene^43^), and *DCAF7* (binds the DYRK1A neurodevelopmental kinase.^44^ TriC also promotes disassembly of the mitotic checkpoint complex^45^, a regulator of neural progenitor division^46^ and regulates BBSome assembly important for primary cilia development.^47^ While heterozygous variants in *TCP1* have recently been linked to intellectual disability and epilepsy^48^, we identified a predicted deleterious homozygous variant (p.A453E) that segregated with disease in affected siblings in family F776, located within the largest autozygous region (**Figure 5a**). All three affected siblings presented with spastic-ataxic CP (**Supplementary Video 1**), intellectual disability, and ophthalmoplegia.

**Figure 5.**
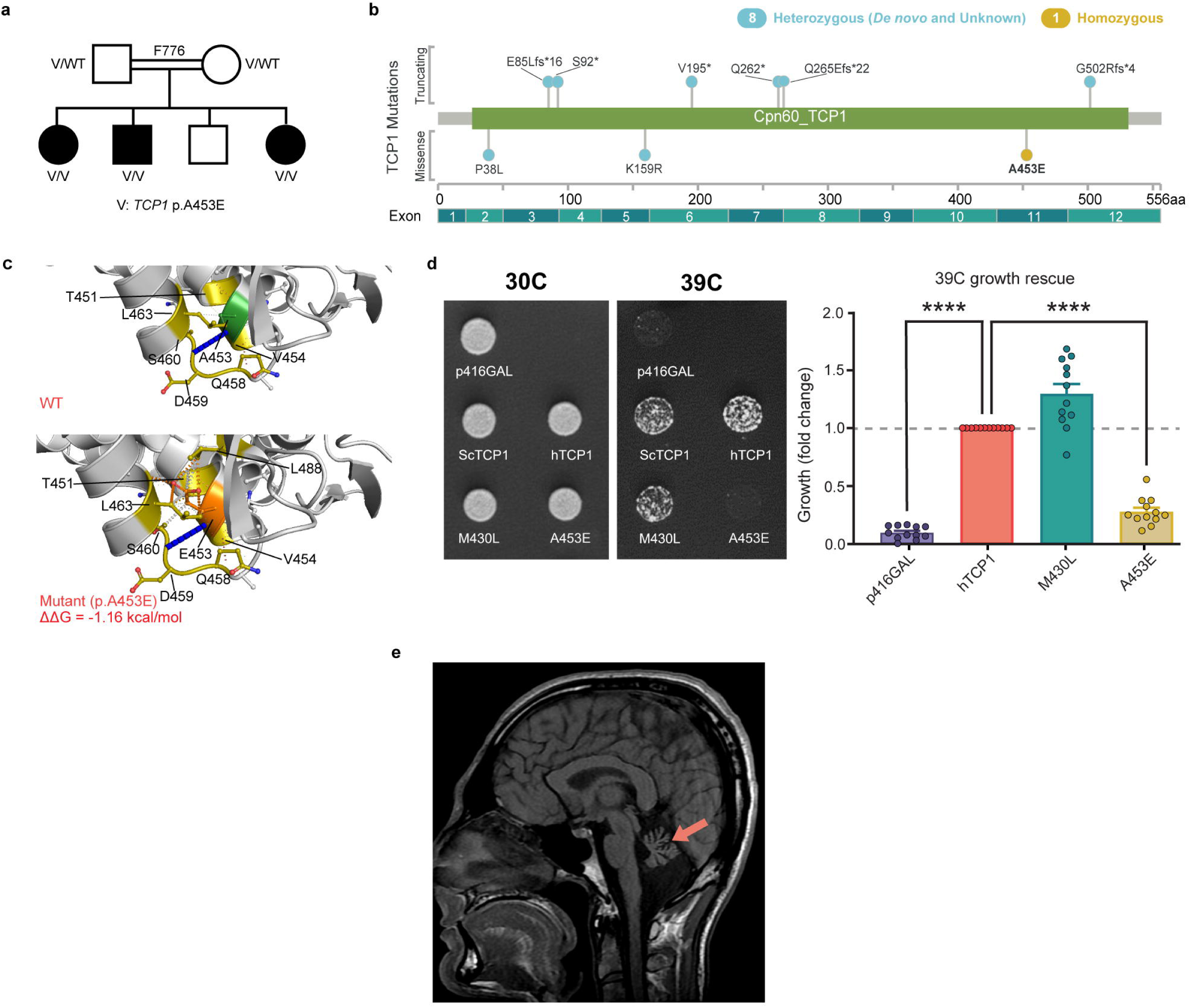
Homozygous TCP1 A453E variants lead to a recessive form of ataxic CP. **a.** TCP1 pedigree show three affected siblings demonstrating compatible phenotypes. **b.** Lollipop plot of TCP1 variants mapped across the T-complex protein 1 subunit alpha, including previously published heterozygous variants.^46^ **c.** DynaMut2 predictions of T-complex protein 1 subunit alpha due to missense variant in the TCP1 gene (p.A453E). N, O, S atoms are represented in blue, red, and yellow, respectively. Hydrophobic, hydrogen-bonding, amide, ionic, and Van der Waal’s (VdW) interactions are shown in green, red, blue, yellow, and light blue, respectively. Top: wild-type (WT) protein structure as predicted by AlphaFold v2 (UniProt accession: P17987). Bottom: mutant protein structure with substituted residue. WT residue is in green, and the substituted residue is in orange. The corresponding calculated ΔΔG value is −1.16 kcal/mo **d.** Functional studies were performed by introducing wild-type yeast tcp1 (scTCP1), human reference TCP1 (hTCP1), or individual variant-containing TCP1 sequence (M430L and A453E) into a p416GAL URA3 backbone and transforming into temperature sensitive yeast strain tcp1-1 (MATa tcp1-1::KanR his3D1 leu2D0 ura3D0 met15D0). Growth was scored after 3 days incubation at 30°C or 39°C. On the left are representative plate images. Densitometric analysis of three individual transformations are shown to the right. Significance was assessed by one-sided t-test; * indicates p<0.05). **e.** Review of TCP1 brain MRI features show hypoplasia of the vermis and cerebellar hemispheres.

The p.A453E variant identified in family F776 resides within the C-terminal equatorial ATP-binding domain, while previously described heterozygous missense variants acting in a haploinsufficient (p.P38L) or dominant negative fashion (p.K159R) are found in N-terminal portions of the protein (**Figure 5b**). Interestingly, although all previously described *TCP1* disease-associated variants have been heterozygous, both *TCP1* p.A453E heterozygous carrier parents were neurologically healthy. Normal mode analysis and protein modeling showed that the mutant variant (p.A453E) causes multiple steric clashes with nearby amino acid residues which do not occur in the wild-type protein. The resulting loss in free energy of stability is −1.16 kcal/mol indicating a destabilizing variant (**Figure 5c**). Although no patient-derived cells were available, functional complementation assays demonstrated that the *TCP1* p.A453E variant led to loss of function (**Figure 5d**). Brain MRI revealed cerebellar hypoplasia (**Figure 5e**). The three affected siblings showed consistent dysmorphic facial features - square face, broad medial eyebrows, high nasal root, long nose, slightly underdeveloped nasal alae, hanging columella, deep philtrum, thin upper lip, horizontal chin crease, and broad chin (photos available on request).

### *SUCO* as a novel CP-associated gene that can also lead to osteogenesis imperfecta

The SUN domain-containing ossification factor *SUCO* (OPT, SLP1) encodes a widely expressed secretory pathway mediator found at rough endoplasmic reticulum sites.^49^ A patient with biallelic *SUCO* variants and a lethal neonatal skeletal dysplasia was previously published^50^ and heterozygous variants were reported in three patients with mesial temporal lobe epilepsy.^51^ *SUCO* knockout (KO) (*Opt ^-/-^*) mice display impaired bone formation and spontaneous fractures but also had evidence of a neurological phenotype with imbalanced gait and an impaired righting response.^49^ Approximately half of *SUCO* KO animals develop neonatal respiratory distress associated with an underdeveloped ribcage and early neonatal mortality similar to the only prior reported human phenotype. We identified a homozygous missense variant (p.R942T) in *SUCO* predicted to be deleterious in family F804, where two affected siblings presented with symptoms of spastic CP together with an osteogenesis imperfecta phenotype. Subsequent matching efforts identified five additional unrelated families harboring biallelic *SUCO* variants (including two homozygous genomic loss of function variants, p.G154Vfs*5 and c.3265+1G>A), presenting with phenotypes that included spasticity, osteogenesis imperfecta, and additional neurodevelopmental features (**Figure 6a, Supplementary Table 10, Supplementary Video 2).** Identified variants included 2 truncating, 3 missense, and 3 canonical splice site variants (**Figure 6b**). Normal mode analysis and protein modeling performed for the three missense variants, p.R942T, p.M453T, p.W608R, predicted that these variants disrupt key structural interactions present within the SUN domain of the wild-type protein, leading to a destabilizing loss of free energy (**Figure 6c**). Patient cells were unavailable, but functional complementation assays in yeast ortholog *slp1* deletion mutants demonstrated that the slp1-M331T (corresponding to the M453T human variant) led to a loss of *SUCO* activity compared to reference protein (**Figure 6d**). We also performed locomotor studies in the invertebrate Drosophila model system using nervous system-specific RNAi knockdown of the *SUCO* ortholog (CG31678), confirming neurological impairments occur independent of skeletal phenotypes (**Figure 6e**). Neuroimaging revealed cerebellar hypoplasia with bilateral white matter hyperintensities in one family, lissencephaly in another, and partial agenesis of the corpus callosum with cerebral white matter T2 hyperintensities in a third family (**Figure 6f**). Shared facial features in human patients (F804) included a drooping facial appearance, lower lid ectropion, and downslanting palpebral fissures (photos available on request).

**Figure 6.**
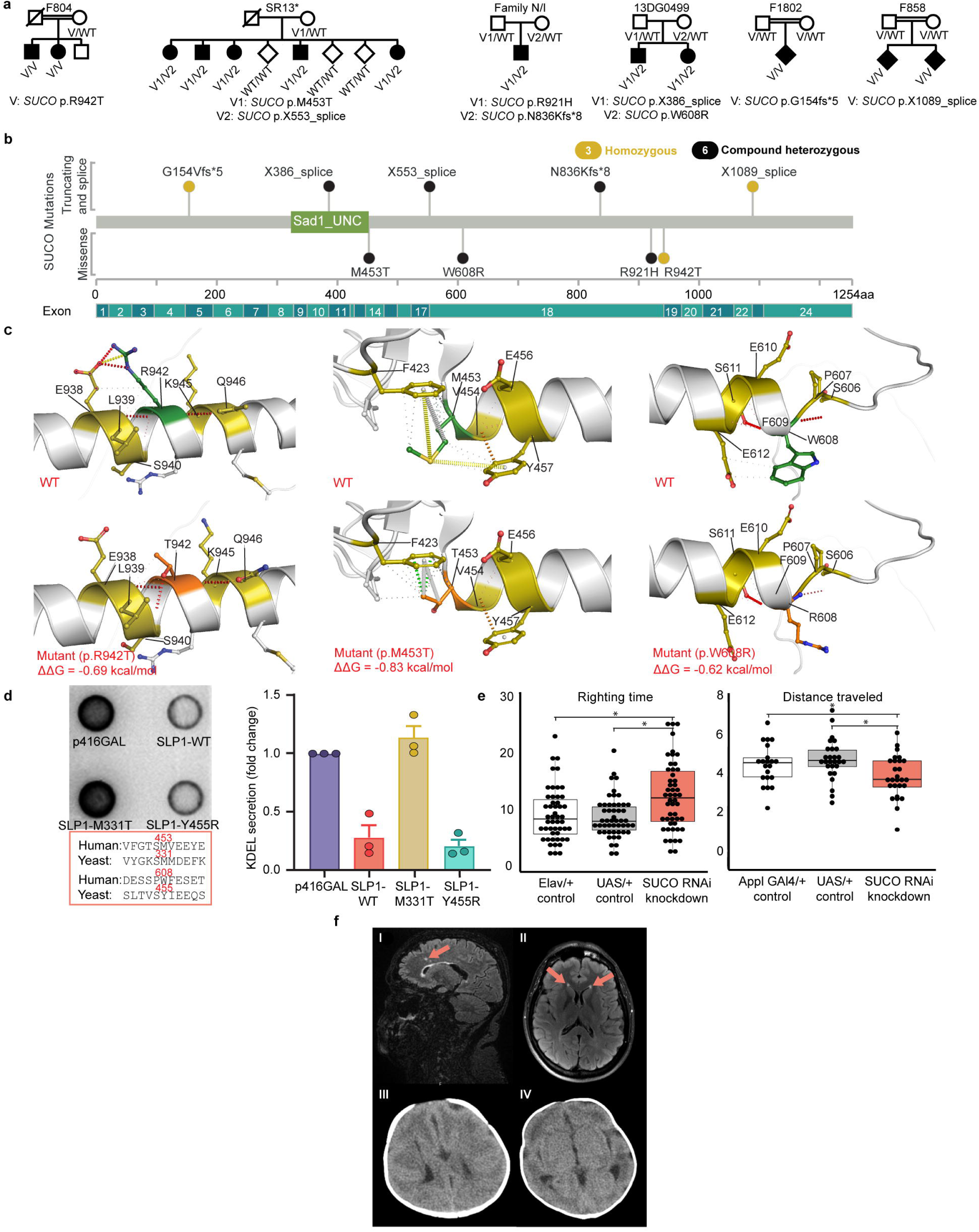
SUCO as a novel CP-associated gene. **a.** SUCO pedigrees: Different variants and inheritance patterns across 6 different families are shown. * indicates partial pedigree. **b.** Lollipop plot of SUCO variants mapped across the SUN domain-containing ossification factor. **c.** DynaMut2 predictions of structural impact on SUN domain-containing ossification factor protein due to missense mutations in SUCO gene (p.M453T, p.W608R, and p.R942T. N, O, S atoms are represented in blue, red, and yellow, respectively. Hydrophobic, hydrogen-bonding, amide, ionic, and Van der Waal’s (VdW) interactions are shown in green, red, blue, yellow, and light blue, respectively. Top: wild-type (WT) protein structure as predicted by AlphaFold v2 (UniProt accession: Q9UBS9). Bottom: mutant protein structure with substituted residue. WT residue is in green, and the substituted residue is in orange. All three substitutions indicated decrease in change of free energy leading to destabilization. The corresponding calculated ΔΔG value is −0.830 kcal/mol using Normal Mode Analysis (NMA). For p.W608R, the corresponding calculated ΔΔG value is −0.622 kcal/mol. For p.R942T, the corresponding calculated ΔΔG value is −0.696 kcal/mol. **d.** Results shows how expression of wild-type SLP1 reduced Kar2p secretion by 70%. The analysis of the variants showed that expression of M331T variant was not able to reduce Kar2p secretion suggesting that specific missense mutation leads to loss of function. However, Y455R variant behaved on a similar way to wild-type SLP1 and was able to decrease Kar2p expression, demonstrating normal functionality for this variant. **e.** Distance traveled in adult locomotor assay. Average time to right in larval turning assay across 3 attempts. *p<0.05 via 2-sided t-test. Box demonstrates 25-75th percentile and median with solid line. Individual trial results plotted (n=20-27 distance, n=50 larval turning). **f.** SUCO brain MRI and CT Scan features. Family SR13 (I) show white matter hyperintensities. Family N/I (II) show bilateral white matter hyperintensities. Family 13DG0499 (III, IV) show lissencephaly.

### Replicating disease associations, expanding phenotypes and inheritance patterns

Based on genotype–phenotype concordance in our cohort, we replicated previously proposed human disease associations for two genes *(SYNRG, SFPQ)* that were detected in a single family or listed in OMIM as having uncertain relationships to monogenic disease **(Supplementary table 11)**, leading to designation as high-confidence candidate genes. Variants in the *NDC1* gene were recently confirmed as a disease-associated gene and published separately.^52^ We also detected variants in *INO80*, previously associated with immunodeficiency with defective immunoglobulin class switching^53^, in the proband of family F1261 with generalized hypotonic CP and intellectual disability. We observed neurological phenotypic expansions in additional five genes (*CACNA1A, TRAPPC9, GRID2, IGHMBP2, DDHD2*), identifying previously unreported clinical manifestations. These expansions included microcephaly, epilepsy, optic nerve atrophy, novel movement disorders, and more severe presentations (**Supplementary Table 12**). In three genes previously reported to follow autosomal dominant inheritance (*TCP1, SPTLC2, TRIO*), we identified homozygous or compound heterozygous variants segregating with CP under an autosomal recessive model. Some of these cases displayed more severe or syndromic phenotypes, highlighting the importance of considering alternative inheritance modes in diverse populations and in clinical interpretation (**Supplementary Table 13**). In addition, previously unreported dysmorphic facial features were observed across seven families representing six different genes (*ALG2, CACNA2D2, POMGNT1, SPATA5, RTTN, SLC1A4*) expanding the human morbid phenotype associated with these monogenic forms of CP (**Table 1**).

**Table 1.**
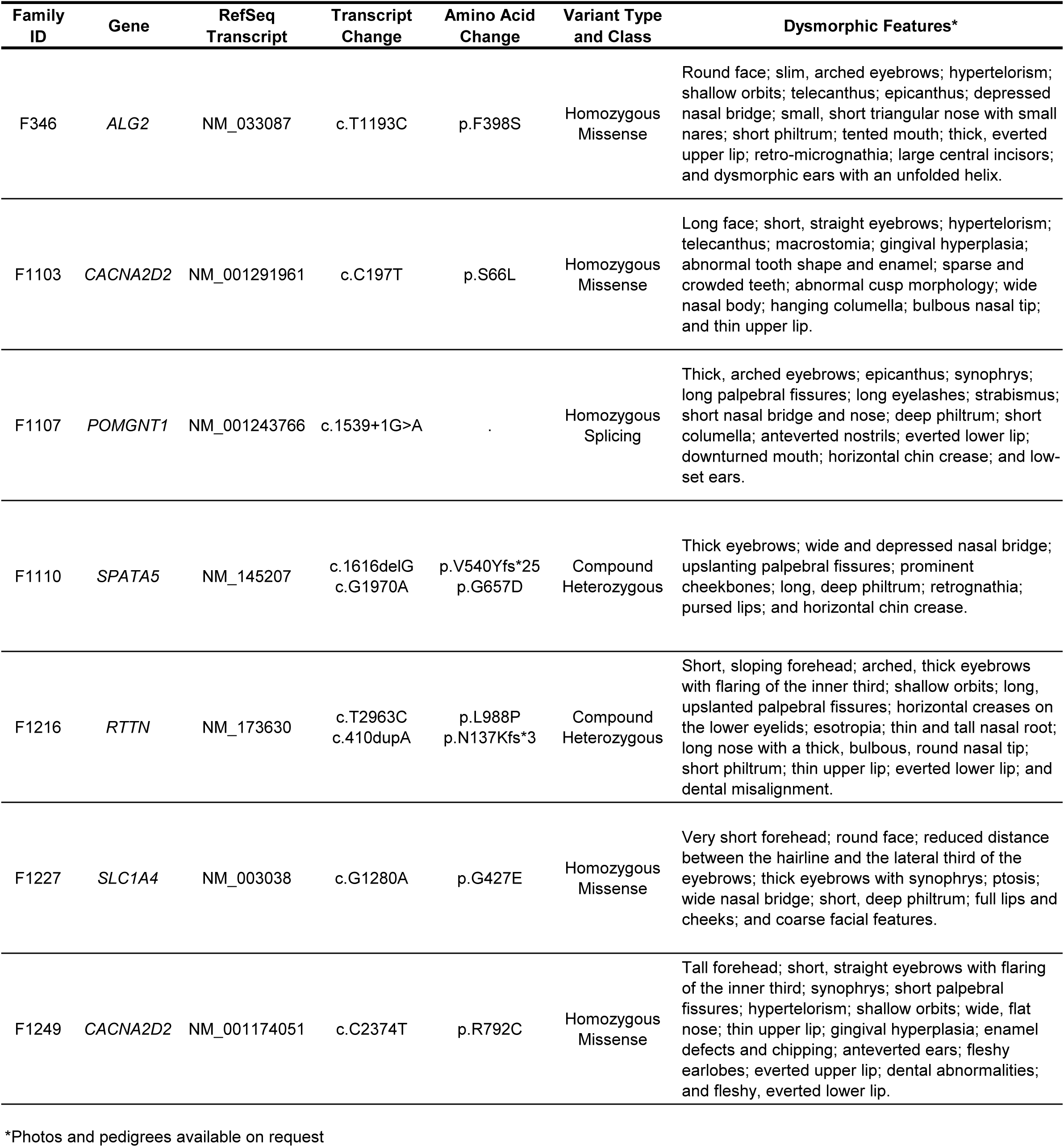
Previously unreported dysmorphic features in affected probands for monogenic forms of CP.

### Diagnostic and therapeutic implications

We observed likely gene-environment interactions in four genes (**Supplementary Table 14**). These represent cases wherein genetic variants have been shown to increase vulnerability to CP-associated acquired insults. For instance, Pathogenic or Likely Pathogenic variants in *G6PD* predispose to kernicterus associated with neonatal jaundice, while *COL4A2* and *ABCC6* are associated with both ischemic and hemorrhagic stroke. Early detection of these variants could guide anticipatory management in high-risk neonates.

34 families (24.82% of our cohort) carried clinically actionable variants, defined as findings with established therapeutic interventions that could impact CP-associated outcomes.^40^ Some of these included examples of dietary interventions (*GK, TXBP1, GAMT, G6PD, PAH, BCKDK, GCDH*), micronutrient supplementation (*KCNJ10, COQ8A, MMACHC*), early screenings and detections *(AH1, KIF7, SIL1, TMEM67, ATM)* and substrate supplementation to reduce toxic metabolite accumulation (*SPTLC2, GAMT*), underscoring the value of genetic diagnosis in guiding early, individualized treatment strategies **(Supplementary Table 15**). These results expand gene-environment context and therapeutic implications for consanguineous populations.

### Gene network analysis of the MAP CP cohort

To assess convergence among CP-associated genes, we integrated gene network, co-expression, and cell-type enrichment analyses. Network clustering of CP-associated genes revealed that subsets of genes co-segregated into clusters based on functional similarities (**Figure 7a**). GO overrepresentation and pathway analysis highlighted a significant enrichment of the AP-4 adaptor complex (GO:0030124), aromatic amino acid metabolism (GO:0009072), glutamate receptor activity (GO:0008066) and phospholipase activity (GO:0004620). KEGG pathway analysis highlighted enrichment of folate biosynthesis (hsa00790), long-term depression (hsa04730), sphingolipid mechanism (hsa00600) and glutamatergic synapses (hsa04724). (**Figure 7b, Supplementary Table 16,17**). Weighted gene co-expression network analysis (WGCNA) was carried out using two distinct gene sets – constraint-based and curated. The analysis of prenatal cortical transcriptomes over these two gene sets identified three modules – blue, midnightblue, and greenyellow – with overrepresentation of CP genes (**Supplementary Figure 4**). Blue and midnightblue were linked to neuronal morphogenesis, while greenyellow (constraint-based) mapped to synaptic signaling and dyskinetic traits (**Supplementary Figure 5**). To extend these analyses to the cellular level, we tested for enrichment of CP-associated genes across midgestational cortical cell types using marker sets derived from single-cell RNA-seq data (17-18 gestational weeks). Logistic regression models accounting for GC content and gene length identified significant overrepresentation of constraint-based and curated gene sets in migrating excitatory neurons (**Figure 7c**), suggesting these cell types contribute to CP risk during cortical development.

**Figure 7.**
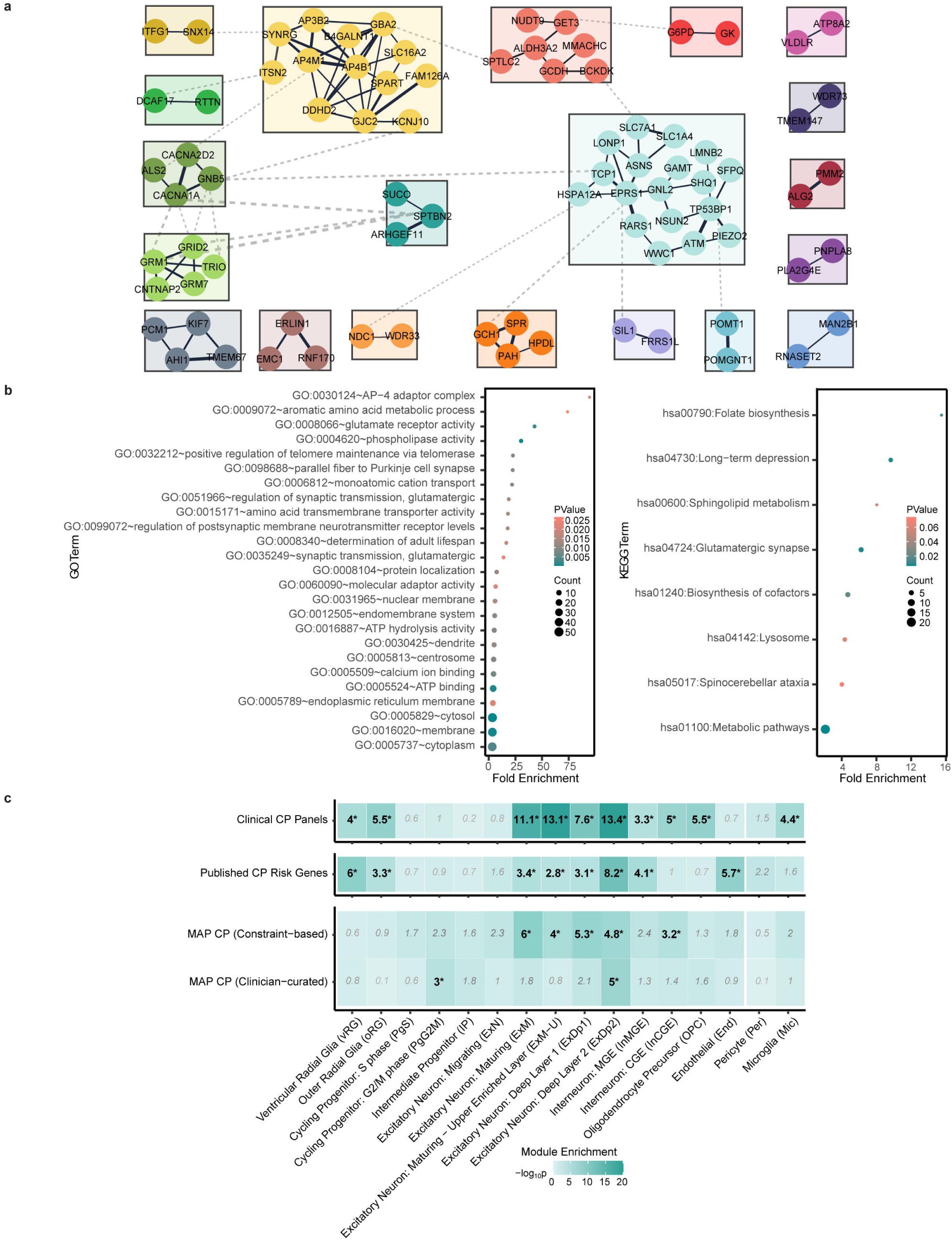
Gene Regulatory Networks, Ontology, and Pathway Analyses. **a.** STRING network cluster of causative homozygous recessive genes identified in the cohort (confidence score = 0.4) features prominent subclusters (indicated by different colors) that were identified on the basis of STRING score similarities using Markov Chain algorithm in Cytoscape. **b.** Enrichment results from gene ontology (GO) and KEGG pathway overrepresentation analyses of causative autozygous and hemizygous recessive genes. Annotations were performed using DAVID. Significance is calculated based on a modified Fischer’s test (EASE). **c.** Enrichment of CP gene sets across prenatal cortical cell types defined by single-cell RNA-seq of 17–18 gestational week cortex. Gene sets tested include clinical panels, published CP risk genes^8^, constraint-based candidates, and clinician-curated genes. Logistic regression was used to assess enrichment, controlling for gene length and GC content. The heatmap shows −log_₁₀_P values; asterisks indicate nominal p<0.05.

## DISCUSSION

Historically, a genetic basis for CP was not suspected given its predominantly sporadic occurrence and implicit bias that CP is an acquired condition.^54,55^ Recent analyses have indicated that more cases of CP may be attributable to genetic causes than to birth-associated injuries.^4,9,10^ Previously published studies, largely focused on CP cohorts from North America, Australia, and mainland China, have found enrichment of damaging *de novo* and transmitted variants that likely contribute to the largely sporadic occurrence of CP.^5–8^ However, these prior studies also found that about one-third of CP-associated genes demonstrated recessive inheritance. Findings from our Middle Eastern cohort diversify our knowledge of recessively-acting CP-associated variants, demonstrating a role for homozygous variants associated with new and complex phenotypes.

Our findings affirm the relevance of prior knowledge of parental consanguinity, particularly in multiplex families, to identify monogenic etiologies of CP but also provide important context. In the MAP CP cohort, comparisons of observed vs. expected ROH highlighted that relatively recent but unrecognized consanguinity can complicate autozygosity mapping as disease-associated variants may be found in ROH regions not predicted by pedigrees. It is to be noted that majority of etiologies we identified display autosomal recessive inheritance, which is in concordant with previous observations that heuristically correlate the presence of consanguinity with higher contributions from recessive variants. However, it does not preclude contributions from other modes of inheritance as we observed in our cohort. Finally, at the present time, it can be challenging to distinguish whether the CP in a given family is due to acquired injury, monogenic or represents an MPV-driven condition which can complicate disease gene discovery, functional validation and clinical interpretation.

Our study achieved an etiologic yield of 72.87% (137/188 families), with nearly 90% of causative genes within families demonstrating autosomal recessive inheritance. This yield is comparable to a recent study of neurogenetic disease in consanguineous families^36^ (72%) and higher than that reported for sporadic congenital heart disease^56^ (13.7%). The high yield here is likely attributable to the application of WES in both affected and unaffected family members in multiplex, consanguineous families, providing robust opportunities for disease-gene characterization through autozygosity mapping and segregation analysis.

We recognized many previously reported CP-associated genes in our cohort, but also identified more than two dozen potential novel CP-gene associations. We found strong support for *CASKIN2*, *SLC7A1*, and *GET3* as high-confidence CP candidates based on integrated protein structural modeling, identified 4 additional high confidence candidates based on concordant LODB findings (*HSPA12A, TP53BP1, WWC1, and MPND*), and replicated prior associations reported in the literature for *SYNRG* and *SFPQ* for a total of 9 high confidence candidates. We also experimentally confirmed *SUCO* as a CP-associated genes. Of relevance to clinical variant interpretation, we found that recessive variants in *TCP1, SPTLC2,* and *TRIO* can lead to severe and/or complex phenotypes in addition to their more frequently observed dominant inheritance patterns. Finally, we found evidence for previously undescribed but consistent facial morphologic features that segregate with affected status for six genes. These findings may be particularly relevant given recent advances in facial morphometry, which can allow variants of uncertain significance to be reclassified as Likely Pathogenic based on the recognition of a consistent facial phenotype.^57^

Our analyses of both known and novel gene networks revealed evidence for additional biological pathways that contribute to CP. In comparison to the Rho family GTPases and actin and tubulin cytoskeleton networks that featured prominently in our analyses of *de novo*-driven pathways, we identified other prominent processes in this cohort. These included enrichment of vesicular transport (i.e., vesicle-mediated transport [node 125], AP-4 adaptor complexes [GO:0030124], and intracellular protein transport [node 183]); long-term depression (hsa04730), shown to be associated with dystonia^58,59^, complex lipid processing [i.e., sphingolipid metabolism [hsa00600], phospholipase activity [GO:0004620], sterol metabolism [node 10] and GPI anchor biosynthesis [node 6] and excitatory cortical neuronal development (i.e. WGCNA, glutamate receptor activity [GO:0008066] and glutamatergic synapses [hsa04724]).

Despite these advances, our study has several limitations. First, although the etiologic yield from whole exome sequencing (72.87%) in consanguineous families was high, we did not assess noncoding variants or complex structural variation. Short-read whole exome sequencing is limited in its ability to detect deep intronic and regulatory variants, repeat expansions, mobile element insertions, copy-number changes, and perform phasing. Whole genome sequencing – especially long read approaches optimized for structural variation and tandem repeats – may resolve additional unsolved cases and further reduce the diagnostic gap. Second, while we observed potential MPVs in 27.8% of families, the extent to which these variants act additively, epistatically, or as driver–modifier pairs to affect phenotypic changes remains unclear. Clarifying these relationships will require functional studies modeling patient alleles in controlled systems, using approaches such as the sequential knock-in/knock-out of variants in isogenic patient-derived induced pluripotent stem cell (iPSC) neural lineages, with readouts from transcriptomics, proteomics, electrophysiology, and quantitative morphometrics. Third, our cohort consisted largely of Middle Eastern consanguineous families. Because variant structures and autozygosity patterns differ across populations, extending analyses to additional consanguineous groups – and to outbred cohorts – will be critical to generalize findings, identify population-specific risk alleles, and refine penetrance estimates. Cross-cohort replication with harmonized phenotyping will reduce any ascertainment bias. Last but not least, although we experimentally validated a subset of genes, patient-derived cells were generally unavailable, and many assays relied on yeast or Drosophila systems. Higher-order models and patient-derived iPSC-based neurons, glia, and organoids may be well-suited to interrogate human-specific mechanisms, cell-type specificity, and dosage effects in the future, particularly given our findings indicating an important role for mid-gestation migrating excitatory cortical neurons in CP.

Our results have important implications for both the diagnosis and treatment of CP. Our findings indicate that additional CP-associated genes still await discovery. As these genes are characterized, we anticipate this will reduce the proportion of exome or genome-negative CP cases undergoing clinical sequencing. Although studies to date have focused on single gene contributions to CP, intriguing gene-environment interactions are emerging^60,61^. In our cohort, we present several examples of gene-environment interactions leading to CP. Future studies will likely need to consider more complex interactions that may contribute to many cases of CP. In addition, although findings from our cohort indicate that all forms of inheritance may contribute to CP, certain variant types may be overrepresented in specific family structures. Our findings indicate that consanguinity can represent both an aid and a challenge to identifying disease-associated genes, and distinguishing monogenic from MPV-driven etiologies will become increasingly important in the future as gene-targeting therapies continue to be developed. Finally, although CP remains a clinical diagnosis^62^, identifying a genetic cause of an individual’s CP can profoundly impact patient care by identifying clinically actionable genetic etiologies.^6,63^ Actionable etiologies, like those we identified in this cohort, can provide treatment options not part of ‘usual care’ for CP, with potentially important impacts on patient outcomes.^63^

## METHODS

### Case cohorts, enrollment, phenotyping, and exclusion criteria

A total of 692 individuals with CP from 188 families were enrolled through protocols approved by the respective ethics committees at the King Faisal Specialist Hospital and Research Centre, University of Khartoum, Semnan University of Medical Sciences, and King Saud University College of Medicine Research Center. Written informed consent was obtained from all participants. The cohort included 143 multiplex families, 32 trios, 3 duos, and 10 singletons. The primary inclusion criterion was a clinical diagnosis of CP confirmed by pediatric neurologists based on the international consensus definition of CP^64^. Individuals were excluded if they had a known acquired risk factor for CP, evidence of a progressive neurological disease, or neurodegeneration. Clinical data were obtained via direct assessments and supplemented by clinical notes and hospital discharge summaries. Comorbidity statuses were also collected where available (**Supplementary Figure 7**). Consanguinity status was based on self-report. Saliva or blood samples were collected from affected individuals and available family members. DNA was extracted using standard protocols with support from the Phoenix Children’s Hospital (PCH) Biorepository and Genomics Labs.

### Control Cohort

A control cohort consisted of 1,789 families from the Simons Simplex Collection, each including one child with autism, one unaffected sibling, and unaffected parents.^65^ For this study, only the unaffected sibling and both parents were analyzed. All control individuals were designated as unaffected by the Simons Simplex Collection. Access to genomic data was granted through the National Institute of Mental Health Data Repository.

### Variant alignment, calling, and filtering

#### Exome sequencing

Most trios were sequenced at the Yale Center for Genome Analysis. Genomic DNA from blood (171 families) or saliva (17 families) underwent exome capture using the IDT xGen Exome v1 (366 samples), v2 (304 samples), v3 (7 samples), v4 (5 samples), or Roche MedExome (10 samples). All samples were subsequently sequenced on an Illumina platform.

#### Variant calling

The exome-sequenced data underwent parallel processing through two independent pipelines at Washington University in St. Louis (WashU) and Phoenix Children’s (PCH).^66^ Sequence reads were aligned to the GRCh38 reference genome using BWA-MEM^67^, followed by duplicate read removal and base quality score recalibration according to the GATK (v4) Best Practices workflow^68^ as previously described with updated annotation databases.^8^ Single-nucleotide variants and small indels were identified using GATK HaplotypeCaller, and variant annotation was performed with ANNOVAR^69^, VEP^70^, dbNSFP^71^ (v4.2c), Bravo^29^ (v8) and gnomAD-genome^26^ (v3.1). Missense variants were classified as deleterious (D-Mis) if predicted to be deleterious by MetaSVM^72^ or if they had a CADD^73,74^ (v1.6) score ≥20. LoF variants included stop-gain, stop-loss, frameshift insertions or deletions, canonical splice-site, or start-loss variants. Non-frameshift indels were also captured. Variants predicted to be either LoF or D-Mis were designated as “damaging”. Variant calls were harmonized between WashU and PCH to ensure consistency prior to downstream analysis. Inheritance model–based classification included DNVs, recessive genotypes, and X-linked variants.

#### Sequence quality control

Sequencing quality was assessed at the sample and cohort levels. Reported sex was verified against genetic sex inferred from X and Y chromosome read depth. Sample identity was confirmed using kinship and relatedness estimates to detect contamination or mislabeling. Coverage statistics required a mean target coverage ≥76× with ≥98% of bases covered at ≥8× read depth. Samples not meeting these thresholds were re-sequenced or excluded from further analysis.

#### Variant filtering

DNVs were called using TrioDenovo^75^ and filtered using stringent quality and frequency thresholds. Variants were retained if they had a minor allele frequency (MAF) ≤ 5 × 10^-4^ in Bravo and gnomAD-genome; a minimum of 10 total reads and 5 alternate allele reads in the proband; and an alternate allele ratio ≥20% (or ≥28% if alternate reads <10). In parents, a minimum depth of 10 reference reads and an alternate allele ratio <3.5% were required. Only exonic or canonical splice-site variants were included.

Rare X-linked hemizygous variants were defined by MAF ≤ 5 × 10^-5^ in Bravo and gnomAD-genome and were required to pass GATK Variant Score Quality Recalibration (VQSR). Variants were required to have ≥8 total reads, with a genotype quality ≥20, mapping quality ≥40, and an alternate allele ratio ≥20% in the proband when ≥10 alternate reads were present, or ≥28% if alternate reads were <10.

Recessive genotypes, including homozygous and compound heterozygous variants, were retained if MAF ≤ 10^-3^ in Bravo and gnomAD-genome. These variants were required to pass GATK VQSR and have ≥8 total reads in the proband. Potentially damaging variants were restricted to LoF variants, D-Mis variants, and non-frameshift indels.

In families with complete trios, *de novo*, homozygous, compound heterozygous, and X-linked variants were analyzed; in incomplete trios, only homozygous and X-linked variants were assessed. Variants were retained if present in affected individuals and absent in unaffected relatives. Variants located in segmental duplication regions were excluded using annotations from ANNOVAR. Additionally, all variants of the cohort were also annotated with allele frequencies (GME_AF) from the Great Middle Eastern (GME) Variome database^30^ using ANNOVAR (table_annovar.pl) and variants with minor allele frequencies greater than 0.01 were excluded. All retained variants were subsequently reviewed in IGV for manual validation. Annotations were also performed using the allele frequencies available from Qatar Precision Health Institute-Qatar Biobank^76^ but no significant matches were derived.

#### Kinship analysis

Kinship estimation was performed using phased variant data for each family. VCF files were processed using BCFTools^77^ (v1.14), with multiallelic sites split and a minor allele frequency threshold of 0.01 applied. Files were then split by chromosome and phased using Beagle^78,79^ (v5.4) with default parameters. Imputation was performed using Beagle’s reference panel based on 3,202 samples, and genetic distance was calculated using the PLINK GRCh38 map file.^80,81^ Phased data were used to compute identity-by-descent (IBD) and homozygosity-by-descent estimates using hap-IBD.^82^ The resulting chromosome-wise files were merged into a single VCF file with BCFTools. Finally, pairwise inbreeding coefficients were estimated using the relatedness2 command from VCFtools (v1.14).^83,84^

### Variant interpretation

Variants surviving filtering were systematically reviewed. Variants were assessed based on deleteriousness, segregation among affected individuals (considering recessive, X-linked, and *de novo* inheritance patterns), and phenotypic consistency. Variants in known disease-associated genes were assigned an ACMG^85^ classification using ClinVar^86^ supplemented by the Franklin platform (https://franklin.genoox.com/) to identify Likely Pathogenic or Pathogenic variants. Genes were subclassified as appropriate into one of the following categories: (1) Known disease-associated gene: listed in OMIM^38^ or supported by recent literature with a phenotype consistent with the case; (2) Multilocus pathogenic variants (MPVs): identification of two or more disease-associated genes compatible with a single proband phenotype; (3) Phenotypic expansion: consistent with known disease-associated gene but with novel clinical features; (4) New inheritance pattern: gene previously associated with a different mode of inheritance (e.g., recessive inheritance pattern in a gene known for dominantly-mediated effects).

Families for whom no known disease-associated gene was identified were then reviewed for possible CP candidate genes. Variants appropriately segregating in probands were ranked based on *in silico* predicted deleteriousness, prioritizing those with predicted genomic LoF (canonical splice, start/stop loss/gain, or frameshift) variants >> MetaSVM prediction of deleteriousness >> CADD ≥24) to integrate segregation, population constraint/allele frequency, and predicted variant impact. We then considered molecular function, spatiotemporal (i.e. developing brain) expression, model organism findings, and molecular network data for final prioritization and used GeneMatcher^87^ to identify additional cases with consistent inheritance and phenotypes in novel candidate genes.

To further evaluate novel candidates, we applied protein structural modeling. Among 24 candidate genes, 18 genes with homozygous missense variants were analyzed using *in silico* modeling to assess structural consequences. Protein sequences and structures were obtained from UniProt^88^ and AlphaFold^89^ (v2), respectively (**Supplementary Table 8**). For proteins without available structures, models were generated using the AlphaFold Server^90^ (v3).

Variant-induced structural effects were assessed using multiple tools: MUpro^91^ predicted effects based on sequence; AlphaMissense^92^, DynaMut2^93^, mCSM^94^, DUET^95^, and DDMut^96^ assessed pathogenicity and/or stability using structure-based machine learning; CSM-Potential^97^ mapped site probability for potential protein-protein interaction and Missense3D^98^ predicted structural disruptions due to amino acid substitutions. All structural renderings were generated using PyMol (v3.1.4.1; The PyMOL Molecular Graphics System, Version 3.0 Schrödinger, LLC).

### ROH and Consanguinity estimates

ROH were identified using AutoMap^99^, which applies a sliding window approach to detect extended regions of homozygosity within each individual of a family and visualized using a custom algorithm. A minimum ROH length of 1 Mb was used as the detection threshold. Homozygous variants identified in probands were mapped to these ROH intervals.

Families were grouped based on reported consanguinity: those reporting consanguinity (n=158) and those denying it (n=26). Total ROH burden was compared between these groups and 56 CP family trios from an independent internal US cohort serving as controls (**Supplementary Table 18**). Mann-Whitney U tests were used to evaluate statistical significance of ROH differences across groups. In a subset of 45 families with available pedigrees, the expected ROH burden was estimated assuming a human genome size of 3,200 Mb and compared to the observed ROH burden (**Supplementary Table 19**). Paired t-tests were used to assess the statistical significance of the difference.

To assess the relationship between ROH and recessive pathogenic variants, we examined whether these variants were located within the top three largest ROH intervals per proband. Additional variants located within smaller ROHs or outside of ROH regions were also noted. Segregation of ROH regions was performed using a custom R script to identify ROH segments exclusive to affected individuals. ROHs shared with unaffected family members were excluded. For families with more than one affected child, overlapping ROHs between affected siblings were retained. Recessive variants located in segregating ROH regions were compared with those identified prior to segregation-based filtering.

### DNV Enrichment Analysis

Observed-to-expected (O/E) enrichment of DNVs was assessed using the R package of denovolyzeR^100^, which applies a gene-specific mutability model derived from large-scale sequencing data^101^. Per-gene mutation expectations were computed using trinucleotide context, gene length, and sequencing coverage. To account for cohort-specific coverage, mutation probability tables were generated separately for case trios and 1,789 control trios. Damaging DNVs were defined as LoF or D-Mis. The expected number of DNVs was calculated by summing the gene-specific mutation probabilities for each functional class, multiplied by the number of probands and a factor of two to account for diploidy. O/E enrichment was evaluated using one-tailed Poisson tests, as implemented in denovolyzeR. O/E ratios and nominal *P-*values were calculated for variant classes and gene sets. For gene set enrichment, the expected mutation probability was computed using genes within the specific set of interest.

Recurrence of DNVs within individual genes was assessed by permutation testing (1 million iterations), in which observed DNVs were randomly redistributed across the genome based on per-gene mutability. Empirical *p*-values were derived from the distribution of recurrent events observed in these simulations.^8,22^ To test for gene-level significance, LoF and D-Mis variants were analyzed separately using one-tailed Poisson tests. A Bonferroni correction was applied to adjust for multiple comparisons across 19,365 genes and two variant categories, resulting in a genome-wide significance threshold of α = 0.05 / (19,365 × 2) ≈ 1.29 × 10^-6^. The lower of the two *p*-values (LoF or D-Mis) was retained per gene.

### Pathway Analysis

Genes harboring recessive variants from the set of identified causative genes were used for protein-protein interaction analysis. Network construction and enrichment data were obtained from STRING and visualized using stringApp (v2.2.0) and yFiles Layout Algorithms (v1.1.5) in Cytoscape (v3.10.3).^102,103^ An edge cutoff of 0.4 (moderate confidence) was applied based on the cumulative STRING score. Following STRING analysis, genes were clustered into subgroups based on similarity in STRING scores using the Markov Cluster Algorithm implemented in clusterMaker^104^ (v2.3.4), with the granularity parameter set to 2.5. Gene-set annotation and overrepresentation analysis were performed on the same gene set using DAVID^105,106^ (**Supplementary Table 16,17**) and ToppFun from the ToppGene^107^ Suite (**Supplementary Table 20**). The background gene list for both tools included all human genes available in their respective databases. In DAVID, statistical significance was evaluated using the EASE score, a modified Fisher’s exact test. ToppFun used a hypergeometric test. A significance threshold of *p*-value < 0.05 was applied for both analyses.

### Functional genomic alignment analysis

The Human Gene Connectome (HGC) version 10.00 was used to generate the distances used in the functional genomic alignment.^108^ Genes used to seed the HGC included 1184 genes from the GeneDx Panel (Cerebral Palsy Xpanded T851) as well as CP-associated genes identified in our cohort, including known genes and high confidence novel candidate genes. We used the APE^109^ (Analysis of Phylogenetics and Evolution; v5.7-1) R package to visualize the circular dendrogram. Selected gene clusters were then analyzed using DAVID^110^ (v2025_1) using GOTERM_BP_DIRECT to identify enriched biological processes. Significant GO terms (Benjamini-adjusted *P* < 0.05) were then collapsed using REVIGO^111^ (v1.8.1) with a semantic similarity threshold of 0.5.

### Multilocus pathogenic variant analysis

A list of known disease-causing genes was obtained from the genemap2.txt file downloaded from the OMIM database on January 27, 2025. Genes identified in this cohort with damaging variants (genomic loss of function or MetaSVM = D or CADD≥24) were mapped to OMIM entries and associated phenotypes. Homozygous variants in recessive genes with phenotypic relevance to CP were retained following manual curation. For each family, we evaluated whether these variants occurred within the same ROH segment. The distribution of families harboring movement disorder–associated genes was visualized using the ggplot2 package in R.

### WGCNA analysis

We performed WGCNA^112^ to evaluate systems-level convergence of CP risk genes in the prenatal human brain. Bulk RNA-seq data from midgestational cortex (14–21 post-conception weeks) were obtained from a previously published transcriptomic atlas.^113^ Co-expression modules were defined as previously described using the WGCNA R package.^113,114^ A total of 18 modules were identified and labeled by color; the unassigned gray module was excluded from enrichment testing.

#### Gene set definition

Genes from the MAP CP cohort were curated using two complementary approaches. First approach incorporated a clinically curated gene set comprising of genes that have been classified as known, novel, or exhibiting phenotypic expansion and/or new inheritance mode, based on concordance with patient presentation (n=137). The second gene set identified genes harboring rare protein-altering variants that passed defined constraint-based thresholds. For missense variants, genes were retained if the variant was predicted to be deleterious, had a CADD score ≥ 24, and was located in a gene with mis-Z ≥ 2. Genes with predicted LoF variants were retained if they occurred in genes with a pLI score ≥ 0.9. In compound heterozygotes, both alleles were required to independently meet these thresholds (n=116).

Control gene sets were generated from two sources. The first included panel genes compiled from PanelApp (363 genes, v1.390), Baylor (419 genes, BG-1300-P419-1), and Invitae (424 genes, Test code: 55004) CP panels, retaining only those with pLI ≥ 0.9 or mis-Z ≥ 2. The second set comprised CP-associated genes from a previously published exome study^8^, lifted to hg38 and annotated with CADD (v1.6). To ensure comparable stringency, retained variants met the same deleteriousness criteria and included only compound heterozygotes with both alleles independently satisfying the thresholds.

#### Module enrichment

WGCNA module gene lists were derived from a bulk RNA-seq atlas of the midgestational human cortex (14–21 gestational weeks)^113^. Modules were labeled by color, with the gray module (containing unassigned genes) excluded from enrichment testing. To evaluate enrichment of MAP CP genes across modules, we applied a logistic regression model: is.disease ∼ is.module + gene covariates (GC content, gene length, and mean expression), as previously described.^113^ The background included all genes assigned to WGCNA modules. Bonferroni correction was applied for 17 modules (α = 0.05 / 17 = 2.94 × 10^-3^), and this threshold was used to define significance.

#### Functional profiling of enriched modules

To characterize the biological relevance of modules showing significant enrichment, we conducted gene ontology (GO) and phenotype ontology (HPO) analyses. GO Biological Process enrichment was performed using the enrichGO function from the clusterProfiler R package, with annotations from org.Hs.eg.db.^115^ Benjamini–Hochberg correction was applied for multiple testing. All genes assigned to WGCNA modules served as the background. Terms were retained if they passed both nominal *p* < 0.05 and FDR *q* < 0.05.

For HPO enrichment, we used the enricher function with TERM2GENE and TERM2NAME dictionaries derived from the "genes_to_phenotype.txt" file (HPO release v2025-05-06). Benjamini–Hochberg correction was applied, and significance was defined as FDR < 0.05. Terms annotated to 100–1,000 genes were retained. For both GO and HPO ontologies, the top 20 terms by adjusted *p*-value were visualized.

#### Cell type enrichment

We assessed MAP CP gene enrichment in prenatal cortical cell types using markers derived from single-cell RNA-seq of 17–18 gestational week cortex.^116^ Logistic regression was applied: is.cell.type ∼ is.gene + GC content + gene length. The background included all genes expressed in ≥3 cells. Bonferroni correction for 16 comparisons yielded a significance threshold of 3.13 × 10^-3^.

### Drosophila rearing and motor control assays

Drosophila melanogaster was reared on standard cornmeal-yeast-sucrose food from the BIO5 media facility (University of Arizona). Stocks were maintained at 25°C under 60–80% relative humidity and a 12 h:12 h light–dark cycle. Control and mutant cultures were reared under identical conditions, including vial density. Mixed-sex cohorts were used in all experiments. To investigate the role of *SUCO*, we used the GAL4-UAS system to knock down its Drosophila ortholog, CG31678 (DIOPT score 12), in postmitotic neurons. The following lines were obtained from the Bloomington Drosophila Stock Center (NIH P40OD018537): Appl-GAL4 (#32040), Elav-GAL4; DCR2-UAS (#25750), UAS-SUCO-RNAi (#56872), w1118 (#3605) backcrossed to Canton-S (#64349) for 12 generations with red-eye selection. Motor control phenotypes were assessed using larval turning and adult locomotor distance assays, as described previously.^8,117^ For the adult distance traveled assay, flies were tested in paired, coded vials. After being tapped to the bottom, a still frame was captured 3 seconds post-tap, and the vertical distance from the midline of each fly to the bottom of the vial was measured using the ImageJ^118^ measure distance function. A total of 27 trials were analyzed. For larval turning, 50 larvae per genotype were flipped onto their dorsal side, and the time required to turn to the ventral surface and initiate forward crawling was recorded for three trials per larva. The average turning time per larva was calculated. Driver selection was phenotype-specific: Elav-GAL4 was used in larval assays to avoid freezing observed in Appl controls, while Appl-GAL4 was used for the adult assay because Elav-driven flies responded weakly to tapping. Statistical comparisons were made using 2-sided Student’s t-test. All plots and analyses were generated in R.

### Yeast complementation assays

#### TCP1 complementation assay

A temperature-sensitive S. cerevisiae strain (tcp1-1: MATa tcp1-1::KanR his3Δ1 leu2Δ0 ura3Δ0 met15Δ0) was kindly provided by the Boone lab (University of Toronto). Human TCP1 transcripts and corresponding variants were codon optimized for yeast and synthesized by Genscript Inc. Wild-type and variant forms were cloned into p416GAL^119^ (URA3 selection marker) using SpeI/SalI restriction sites. Yeast TCP1 was cloned into the same vector using EcoRI/HindIII. Yeast transformations were performed using the lithium acetate protocol.^120^ For growth rescue assays, tcp1-1 transformants were precultured overnight in synthetic defined medium lacking uracil. Equal numbers of cells were spotted onto YPD agar and incubated for 3 days at either 30°C (permissive) or 39°C (restrictive) before growth was documented and scored.

#### SUCO variant assessment via Kar2p secretion assays

Yeast slp1 is an ER membrane protein that forms a complex with Emp65 and binds unfolded ER luminal proteins to shield them from premature ER-associated degradation until productive folding can occur. Loss of Slp1 causes normal luminal clients to be aberrantly degraded, triggering ER stress. The resulting stress is thought to increase production of proteins with an HDEL retrieval tag. More of these proteins are packaged into COPII-positive vesicles and nonspecifically delivered to the Golgi. This then overloads the HDEL-mediated ER retrieval pathway, causing marker proteins such as yeast BiP (Karp2) to enter the secretory pathway. We used the karp2 secretion assay to assess secretory pathway overflow in slp1 mutants and determined the ability of human SUCO reference sequence (wild-type) and variants to rescue this function.

Kar2p features an endogenous C-terminal HDEL ER retrieval tag used as a readout of ER retrieval/secretory pathway integrity; if the ER retrieval pathway is saturated or impaired, Kar2p will instead be nonspecifically secreted into the medium. The *S. cerevisiae* slp1-Δ strain was transformed with wild-type and mutant human *SUCO* (M331T, Y455R) constructs cloned into the p416GAL expression vector. Transformed strains were cultured to saturation in synthetic defined medium lacking uracil (SD-URA), and 1×10^6^ cells were spotted onto yeast peptone dextrose (YPD) agar plates overlaid with nitrocellulose membranes. Plates were incubated at 30°C for 24 hours. Yeast cells were then removed from the membranes by washing under running distilled water. Nitrocellulose membranes were blocked in 5% BSA in Tris Buffered Saline with 0.1% Tween-20 (TTBS) for 1 hour and incubated with HDEL primary antibody (Santa Cruz Biotechnology, sc-53472; 1:5,000 in TTBS, 1 hour), followed by HRP-conjugated anti-mouse secondary antibody (1 hour). Signal was detected via enhanced chemiluminescence. Kar2p secretion signals were normalized to those observed in cells transformed with the empty p416GAL vector.

### Clinical annotation and phenotypic expansion analysis

Comprehensive clinical review and phenotypic annotation were performed for all individuals in the MAP CP cohort. Data were curated from original medical records using REDCap^121,122^ and cross-referenced with established gene-disease databases, including OMIM, ClinVar, and GeneReviews^123^. Clinical evaluations focused on neurological phenotypes, atypical presentations, and assessment of inheritance patterns. For all genes listed in OMIM as having “uncertain significance” or lacking confirmed human disease association, we compiled prior published clinical descriptions and compared them to phenotypes observed in our cohort. Genes previously associated with autosomal dominant inheritance were reassessed in cases where variants appeared in homozygous or compound heterozygous states.

To evaluate actionability, genes identified in this cohort were evaluated as we previously described.^6,63^ Potential gene-environment interactions contributing to CP pathogenesis were assessed via curated literature review.

Dysmorphology was assessed by a trained clinical dysmorphologist (CGM). Frontal facial photographs were collected for patients harboring pathogenic or likely pathogenic variants within STRING-defined gene clusters. Composite facial images were generated using Face2Gene^124^ for visual comparison across functional clusters.

To assess novel phenotypes, dysmorphic features were compared against OMIM entries for each gene and supplemented by primary literature review. Features not previously reported were compiled into summary tables and figures. A genotype-phenotype summary table was generated for all individuals with biallelic pathogenic or likely pathogenic *SUCO* variants, incorporating both internal and external cases^50^.

## DATA AND CODE AVAILABILITY

Sequencing data, patient photos, and videos are available on request.

The softwares utilized in this study is available at the following web addresses:

FastQC v0.12.1 (https://www.bioinformatics.babraham.ac.uk/projects/fastqc/)

MultiQC v1.14 (https://github.com/MultiQC/MultiQC)

GATK v4.2.6.1 (https://gatk.broadinstitute.org/hc/en-us/sections/5358821689883-4-2-6-1)

ANNOVAR v4.2 (Date:2020-06-07) (https://annovar.openbioinformatics.org/en/latest/)

VEP release 114 (https://useast.ensembl.org/info/docs/tools/vep/index.html)

VCFtools v0.1.16 (https://github.com/vcftools/vcftools/releases/tag/v0.1.16)

BWA v0.7.17 (https://github.com/lh3/bwa/releases/tag/v0.7.17)

Samtools v1.16.1 (https://github.com/samtools/samtools/releases/tag/1.16.1)

Picard v2.27.5 (https://github.com/broadinstitute/picard/releases/tag/2.27.5)

IGV v2.16.1 (Linux) (https://igv.org/doc/desktop/)

Bedtools v2.30.0 (https://github.com/arq5x/bedtools2/releases/tag/v2.30.0)

TrioDenovo v0.06 (https://genome.sph.umich.edu/w/index.php?title=Triodenovo&section=0&mobileaction=toggle_view_desktop)

R v4.3.3 (https://www.R-project.org/)

Python v3.11.5 (https://www.python.org/)

PLINK v1.9 https://www.cog-genomics.org/plink/)

Face2Gene (https://www.face2gene.com)

MutationMapper (https://www.cbioportal.org/mutation_mapper)

stringApp v2.2.0 (https://apps.cytoscape.org/apps/stringapp)

clusterMaker v2.3.4 (https://apps.cytoscape.org/apps/clustermaker2)

Franklin v76.1+ (https://franklin.genoox.com/)

DAVID 2021 (updated v2023q4) (https://davidbioinformatics.nih.gov)

ToppGene (updated Nov 2024) (https://toppgene.cchmc.org)

REVIGO v1.8.1 (https://github.com/rajko-horvat/RevigoWeb)

Beagle v5.4 (https://faculty.washington.edu/browning/beagle/b5_4.html)

VCFtools v1.14 (https://github.com/vcftools/vcftools/releases/tag/v0.1.14)

BCFTools v1.14 (https://github.com/samtools/bcftools/releases/tag/1.14) ImageJ (https://imagej.net/ij/)

Scripts used in this analysis are available at https://github.com/Kruer-Lab/MAPCP/tree/main.

The GATK pipeline used for calling variants is available at https://github.com/Kruer-Lab/wes_gatk_v2

## ETHICS STATEMENT

This study was approved by the Institutional Review Board of Phoenix Children’s. All participants have provided written informed consent prior to participation. For privacy protection, all participant families have been assigned and referenced using project-specific family identifiers that are de-identified and accessible only to the research team. These identifiers are unique and cannot be linked to individual identities involved in the study.

## Data Availability

All data produced in the present study are available upon reasonable request to the authors

https://github.com/Kruer-Lab/wes_gatk_v2

https://github.com/Kruer-Lab/MAPCP/tree/main

## ACKNOWLEDGMENTS

MCK is supported by NIH R01NS106298 and NIH R01NS127108. SCJ is supported by the Cerebral Palsy Alliance Research Foundation Project Grant (PRG03121), WashU Children’s Discovery Institute Faculty Scholar award (CDI-FR-2021-926), and NIH R01NS131610.

## AUTHOR INFORMATION

MCK serves on the Scientific Advisory Board for United Cerebral Palsy, Merz, and Aeglea. MCK is a consultant for Acadia, BridgeBio, Neurocrine, PTC Therapeutics. MCK also receives grant funding from Medtronic and PTC Therapeutics. All other authors have no declarations of interest.

## CONTRIBUTIONS

M.C.K., S.C.J., H.D., F.A., Y-C.W, P.B., S.B., P.T.S, S.L., S.P-L., H.M., H.Z., H.S., S.S., F.M. contributed to study design, data interpretation and oversight.

H.D, H.M, C.I.G-M, M.C.K., J.L., I.N.M., M.A.S, S.B., A.R., S.G.F., S.S., A.T., E.T., E.N., S.V., J.J., S.A., S.A.H., A.K., F.J., A.A., H.K., P.D., F.A., B.A., K.B., S.M., M.O.H. provided cohort ascertainment, recruitment, and phenotypic characterization.

Y-C.W, S.B., P.T.S., P.B., N.K., Y.X., W.Z., B.L., H.Z., T.N.K, F.V.R. performed genomic and statistical analyses.

P.B. performed proteomic structural analyses.

Y-C.W., P.B, P.T.S. performed pathway analyses.

S.P.M., J.D., M.S., J.L.L., R.R., G.S., H.S., M.B., B.D., N.H., J.B. provided additional cases for *SUCO* gene reporting.

S.L. performed *Drosophila* locomotor experiments. S.P-L performed *Yeast* complementation assays.

P.B., Y-C.W., P.T.S., C.I.G-M., generated figures, tables, and supplementary files.

Y-C.W., P.B., P.T.S., P.T.S. C.I.G-M., S.B., S.C.J., M.C.K. wrote the initial manuscript

H.D., M.C.K., S.C.J, F.A., S.L., M.S., H.S., B.D., J.L.L, F.M., J.D., M.A.S., M.O.H, H.Z. reviewed and edited the manuscript

H.D., S.C.J., M.C.K. acquired funding and supervised the project and were considered co-senior authors

All authors have read and approved the final manuscript.

## REFERENCES

1. Yeargin-Allsopp, M. et al. Prevalence of cerebral palsy in 8-year-old children in three areas of the United States in 2002: a multisite collaboration. Pediatrics 121, 547–54 (2008).

2. Graham, H.K. et al. Cerebral palsy. Nat Rev Dis Primers 2, 15082 (2016).

3. Smith, S.E. et al. Adults with Cerebral Palsy Require Ongoing Neurologic Care: A Systematic Review. Ann Neurol 89, 860–871 (2021).

4. Nelson, K.B. & Blair, E. Prenatal Factors in Singletons with Cerebral Palsy Born at or near Term. N Engl J Med 373, 946–53 (2015).

5. Fehlings, D.L. et al. Comprehensive whole-genome sequence analyses provide insights into the genomic architecture of cerebral palsy. Nat Genet 56, 585–594 (2024).

6. Wang, Y. et al. Exome sequencing reveals genetic heterogeneity and clinically actionable findings in children with cerebral palsy. Nat Med 30, 1395–1405 (2024).

7. McMichael, G. et al. Whole-exome sequencing points to considerable genetic heterogeneity of cerebral palsy. Mol Psychiatry 20, 176–82 (2015).

8. Jin, S.C. et al. Mutations disrupting neuritogenesis genes confer risk for cerebral palsy. Nat Genet 52, 1046–1056 (2020).

9. Srivastava, S. et al. Molecular Diagnostic Yield of Exome Sequencing and Chromosomal Microarray in Cerebral Palsy: A Systematic Review and Meta-analysis. JAMA Neurol 79, 1287–1295 (2022).

10. Gonzalez-Mantilla, P.J. et al. Diagnostic Yield of Exome Sequencing in Cerebral Palsy and Implications for Genetic Testing Guidelines: A Systematic Review and Meta-analysis. JAMA Pediatr 177, 472–478 (2023).

11. Srivastava, S. et al. Meta-analysis and multidisciplinary consensus statement: exome sequencing is a first-tier clinical diagnostic test for individuals with neurodevelopmental disorders. Genet Med 21, 2413–2421 (2019).

12. Janzing, A.M., Eklund, E., De Koning, T.J. & Eggink, H. Clinical Characteristics Suggestive of a Genetic Cause in Cerebral Palsy: A Systematic Review. Pediatr Neurol 153, 144–151 (2024).

13. Moreno-De-Luca, A. et al. Molecular Diagnostic Yield of Exome Sequencing in Patients With Cerebral Palsy. JAMA 325, 467–475 (2021).

14. Modell, B. & Darr, A. Science and society: genetic counselling and customary consanguineous marriage. Nat Rev Genet 3, 225–9 (2002).

15. Albanghali, M.A. Prevalence of Consanguineous Marriage among Saudi Citizens of Albaha, a Cross-Sectional Study. Int J Environ Res Public Health 20(2023).

16. Hosseini-Chavoshi, M., Abbasi-Shavazi, M.J. & Bittles, A.H. Consanguineous marriage, reproductive behaviour and postnatal mortality in contemporary Iran. Hum Hered 77, 16–25 (2014).

17. Hussein, W.M. et al. Correlates and reproductive consequences of consanguinity in six Egyptian governorates. Afr J Reprod Health 26, 48–56 (2022).

18. al-Gazali, L.I., et al. Consanguineous marriages in the United Arab Emirates. J Biosoc Sci 29, 491–7 (1997).

19. Koc, I. & Eryurt, M.A. The Causal Relationship between Consanguineous Marriages and Infant Mortality in Turkey. J Biosoc Sci 49, 536–555 (2017).

20. Sinha, G., Corry, P., Subesinghe, D., Wild, J. & Levene, M.I. Prevalence and type of cerebral palsy in a British ethnic community: the role of consanguinity. Dev Med Child Neurol 39, 259–62 (1997).

21. Daher, S. & El-Khairy, L. Association of cerebral palsy with consanguineous parents and other risk factors in a Palestinian population. East Mediterr Health J 20, 459–68 (2014).

22. Jin, S.C. et al. Contribution of rare inherited and de novo variants in 2,871 congenital heart disease probands. Nat Genet 49, 1593–1601 (2017).

23. McManus, V., Guillem, P., Surman, G. & Cans, C. SCPE work, standardization and definition--an overview of the activities of SCPE: a collaboration of European CP registers. Zhongguo Dang Dai Er Ke Za Zhi 8, 261–5 (2006).

24. Smithers-Sheedy, H. et al. A special supplement: findings from the Australian Cerebral Palsy Register, birth years 1993 to 2006. Dev Med Child Neurol 58 Suppl 2, 5–10 (2016).

25. Karczewski, K.J. et al. The mutational constraint spectrum quantified from variation in 141,456 humans. Nature 581, 434–443 (2020).

26. Chen, S. et al. A genomic mutational constraint map using variation in 76,156 human genomes. Nature 625, 92–100 (2024).

27. Lek, M. et al. Analysis of protein-coding genetic variation in 60,706 humans. Nature 536, 285–91 (2016).

28. Karczewski, K.J. et al. The ExAC browser: displaying reference data information from over 60 000 exomes. Nucleic Acids Res 45, D840–D845 (2017).

29. Taliun, D. et al. Sequencing of 53,831 diverse genomes from the NHLBI TOPMed Program. Nature 590, 290–299 (2021).

30. Scott, E.M. et al. Characterization of Greater Middle Eastern genetic variation for enhanced disease gene discovery. Nat Genet 48, 1071–6 (2016).

31. Alazami, A.M. et al. Accelerating novel candidate gene discovery in neurogenetic disorders via whole-exome sequencing of prescreened multiplex consanguineous families. Cell Rep 10, 148–61 (2015).

32. Reuter, M.S. et al. Diagnostic Yield and Novel Candidate Genes by Exome Sequencing in 152 Consanguineous Families With Neurodevelopmental Disorders. JAMA Psychiatry 74, 293–299 (2017).

33. Anazi, S. et al. Clinical genomics expands the morbid genome of intellectual disability and offers a high diagnostic yield. Mol Psychiatry 22, 615–624 (2017).

34. Wakeling, M.N. et al. Homozygosity mapping provides supporting evidence of pathogenicity in recessive Mendelian disease. Genet Med 21, 982–986 (2019).

35. Pemberton, T.J. et al. Genomic patterns of homozygosity in worldwide human populations. Am J Hum Genet 91, 275–92 (2012).

36. Hiz Kurul, S., et al. High diagnostic rate of trio exome sequencing in consanguineous families with neurogenetic diseases. Brain 145, 1507–1518 (2022).

37. Harrison, S.M., Biesecker, L.G. & Rehm, H.L. Overview of Specifications to the ACMG/AMP Variant Interpretation Guidelines. Curr Protoc Hum Genet 103, e93 (2019).

38. Amberger, J.S., Bocchini, C.A., Schiettecatte, F., Scott, A.F. & Hamosh, A. OMIM.org: Online Mendelian Inheritance in Man (OMIM(R)), an online catalog of human genes and genetic disorders. Nucleic Acids Res 43, D789–98 (2015).

39. Posey, J.E. et al. Resolution of Disease Phenotypes Resulting from Multilocus Genomic Variation. N Engl J Med 376, 21–31 (2017).

40. Mitani, T. et al. High prevalence of multilocus pathogenic variation in neurodevelopmental disorders in the Turkish population. Am J Hum Genet 108, 1981–2005 (2021).

41. Monies, D. et al. Lessons Learned from Large-Scale, First-Tier Clinical Exome Sequencing in a Highly Consanguineous Population. Am J Hum Genet 104, 1182–1201 (2019).

42. Bakur, K. et al. Adult genomic medicine: lessons from a multisite study of 2700 patients. Genome Med 17, 105 (2025).

43. Choi, H.B. et al. Case report: Suspecting guanine nucleotide-binding protein beta 1 mutation in dyskinetic cerebral palsy is important. Front Pediatr 11, 1204360 (2023).

44. Glenewinkel, F. et al. The adaptor protein DCAF7 mediates the interaction of the adenovirus E1A oncoprotein with the protein kinases DYRK1A and HIPK2. Sci Rep 6, 28241 (2016).

45. Kaisari, S., Sitry-Shevah, D., Miniowitz-Shemtov, S., Teichner, A. & Hershko, A. Role of CCT chaperonin in the disassembly of mitotic checkpoint complexes. Proc Natl Acad Sci U S A 114, 956–961 (2017).

46. Carvalhal, S. et al. Biallelic BUB1 mutations cause microcephaly, developmental delay, and variable effects on cohesion and chromosome segregation. Sci Adv 8, eabk0114 (2022).

47. Seo, S. et al. BBS6, BBS10, and BBS12 form a complex with CCT/TRiC family chaperonins and mediate BBSome assembly. Proc Natl Acad Sci U S A 107, 1488–93 (2010).

48. Kraft, F. et al. Brain malformations and seizures by impaired chaperonin function of TRiC. Science 386, 516–525 (2024).

49. Sohaskey, M.L. et al. Osteopotentia regulates osteoblast maturation, bone formation, and skeletal integrity in mice. J Cell Biol 189, 511–25 (2010).

50. Maddirevula, S. et al. Expanding the phenome and variome of skeletal dysplasia. Genet Med 20, 1609–1616 (2018).

51. Sha, Z. et al. Exome sequencing identifies SUCO mutations in mesial temporal lobe epilepsy. Neurosci Lett 591, 149–154 (2015).

52. Smits, D.J. et al. Biallelic NDC1 variants that interfere with ALADIN binding are associated with neuropathy and triple A-like syndrome. HGG Adv 5, 100327 (2024).

53. Kracker, S. et al. An inherited immunoglobulin class-switch recombination deficiency associated with a defect in the INO80 chromatin remodeling complex. J Allergy Clin Immunol 135, 998–1007 e6 (2015).

54. Hemminki, K., Li, X., Sundquist, K. & Sundquist, J. High familial risks for cerebral palsy implicate partial heritable aetiology. Paediatr Perinat Epidemiol 21, 235–41 (2007).

55. Little, W.J. The classic: Hospital for the cure of deformities: course of lectures on the deformities of the human frame. 1843. Clin Orthop Relat Res 470, 1252–6 (2012).

56. Dong, W. et al. Mutation spectrum of congenital heart disease in a consanguineous Turkish population. Mol Genet Genomic Med 10, e1944 (2022).

57. Dingemans, A.J.M. et al. PhenoScore quantifies phenotypic variation for rare genetic diseases by combining facial analysis with other clinical features using a machine-learning framework. Nat Genet 55, 1598–1607 (2023).

58. Martella, G. et al. Impairment of bidirectional synaptic plasticity in the striatum of a mouse model of DYT1 dystonia: role of endogenous acetylcholine. Brain 132, 2336–49 (2009).

59. Lewis, S.A., et al. eIF2alpha phosphorylation evokes dystonia-like movements with D2- receptor and cholinergic origin and abnormal neuronal connectivity. bioRxiv (2024).

60. Massaro, A.N. et al. Whole genome methylation and transcriptome analyses to identify risk for cerebral palsy (CP) in extremely low gestational age neonates (ELGAN). Sci Rep 11, 5305 (2021).

61. Lewis, S.A. et al. AGAP1-associated endolysosomal trafficking abnormalities link gene- environment interactions in neurodevelopmental disorders. Dis Model Mech 16(2023).

62. Aravamuthan, B.R. et al. Uncertainties Regarding Cerebral Palsy Diagnosis: Opportunities to Clarify the Consensus Definition. Neurol Clin Pract 14, e200353 (2024).

63. Lewis, S.A. et al. Clinical Actionability of Genetic Findings in Cerebral Palsy: A Systematic Review and Meta-Analysis. JAMA Pediatr 179, 137–44 (2025).

64. Rosenbaum, P. et al. A report: the definition and classification of cerebral palsy April 2006. Dev Med Child Neurol Suppl 109, 8–14 (2007).

65. Krumm, N. et al. Excess of rare, inherited truncating mutations in autism. Nat Genet 47, 582–8 (2015).

66. Jennewein, D.M. et al. The Sol Supercomputer at Arizona State University. in Practice and Experience in Advanced Research Computing 2023: Computing for the Common Good 296–301 (Association for Computing Machinery, Portland, OR, USA, 2023).

67. Li, H. Aligning sequence reads, clone sequences and assembly contigs with BWA-MEM. arXiv (2013).

68. Van der Auwera, G.A. et al. From FastQ data to high confidence variant calls: the Genome Analysis Toolkit best practices pipeline. Curr Protoc Bioinformatics 43, 11 10 1–11 10 33 (2013).

69. Wang, K., Li, M. & Hakonarson, H. ANNOVAR: functional annotation of genetic variants from high-throughput sequencing data. Nucleic Acids Res 38, e164 (2010).

70. McLaren, W. et al. The Ensembl Variant Effect Predictor. Genome Biol 17, 122 (2016).

71. Liu, X., Li, C., Mou, C., Dong, Y. & Tu, Y. dbNSFP v4: a comprehensive database of transcript-specific functional predictions and annotations for human nonsynonymous and splice-site SNVs. Genome Med 12, 103 (2020).

72. Dong, C. et al. Comparison and integration of deleteriousness prediction methods for nonsynonymous SNVs in whole exome sequencing studies. Hum Mol Genet 24, 2125–37 (2015).

73. Rentzsch, P., Schubach, M., Shendure, J. & Kircher, M. CADD-Splice-improving genome-wide variant effect prediction using deep learning-derived splice scores. Genome Med 13, 31 (2021).

74. Rentzsch, P., Witten, D., Cooper, G.M., Shendure, J. & Kircher, M. CADD: predicting the deleteriousness of variants throughout the human genome. Nucleic Acids Res 47, D886–D894 (2019).

75. Wei, Q. et al. A Bayesian framework for de novo mutation calling in parents-offspring trios. Bioinformatics 31, 1375–81 (2015).

76. Al Thani, A. et al. Qatar Biobank Cohort Study: Study Design and First Results. Am J Epidemiol 188, 1420–1433 (2019).

77. Danecek, P. et al. Twelve years of SAMtools and BCFtools. Gigascience 10(2021).

78. Browning, B.L., Tian, X., Zhou, Y. & Browning, S.R. Fast two-stage phasing of large-scale sequence data. Am J Hum Genet 108, 1880–1890 (2021).

79. Browning, B.L., Zhou, Y. & Browning, S.R. A One-Penny Imputed Genome from Next-Generation Reference Panels. Am J Hum Genet 103, 338–348 (2018).

80. Fairley, S., Lowy-Gallego, E., Perry, E. & Flicek, P. The International Genome Sample Resource (IGSR) collection of open human genomic variation resources. Nucleic Acids Res 48, D941–D947 (2020).

81. Byrska-Bishop, M. et al. High-coverage whole-genome sequencing of the expanded 1000 Genomes Project cohort including 602 trios. Cell 185, 3426–3440 e19 (2022).

82. Zhou, Y., Browning, S.R. & Browning, B.L. A Fast and Simple Method for Detecting Identity-by-Descent Segments in Large-Scale Data. Am J Hum Genet 106, 426–437 (2020).

83. Manichaikul, A. et al. Robust relationship inference in genome-wide association studies. Bioinformatics 26, 2867–73 (2010).

84. Danecek, P. et al. The variant call format and VCFtools. Bioinformatics 27, 2156–8 (2011).

85. Richards, S. et al. Standards and guidelines for the interpretation of sequence variants: a joint consensus recommendation of the American College of Medical Genetics and Genomics and the Association for Molecular Pathology. Genet Med 17, 405–24 (2015).

86. Landrum, M.J. et al. ClinVar: updates to support classifications of both germline and somatic variants. Nucleic Acids Res 53, D1313–D1321 (2025).

87. Sobreira, N., Schiettecatte, F., Valle, D. & Hamosh, A. GeneMatcher: a matching tool for connecting investigators with an interest in the same gene. Hum Mutat 36, 928–30 (2015).

88. UniProt, C. UniProt: the Universal Protein Knowledgebase in 2023. Nucleic Acids Res 51, D523–D531 (2023).

89. Jumper, J. et al. Highly accurate protein structure prediction with AlphaFold. Nature 596, 583–589 (2021).

90. Abramson, J. et al. Accurate structure prediction of biomolecular interactions with AlphaFold 3. Nature 630, 493–500 (2024).

91. Cheng, J., Randall, A. & Baldi, P. Prediction of protein stability changes for single-site mutations using support vector machines. Proteins 62, 1125–32 (2006).

92. Cheng, J. et al. Accurate proteome-wide missense variant effect prediction with AlphaMissense. Science 381, eadg7492 (2023).

93. Rodrigues, C.H.M., Pires, D.E.V. & Ascher, D.B. DynaMut2: Assessing changes in stability and flexibility upon single and multiple point missense mutations. Protein Sci 30, 60–69 (2021).

94. Pires, D.E., Ascher, D.B. & Blundell, T.L. mCSM: predicting the effects of mutations in proteins using graph-based signatures. Bioinformatics 30, 335–42 (2014).

95. Pires, D.E., Ascher, D.B. & Blundell, T.L. DUET: a server for predicting effects of mutations on protein stability using an integrated computational approach. Nucleic Acids Res 42, W314–9 (2014).

96. Zhou, Y., Pan, Q., Pires, D.E.V., Rodrigues, C.H.M. & Ascher, D.B. DDMut: predicting effects of mutations on protein stability using deep learning. Nucleic Acids Res 51, W122–W128 (2023).

97. Rodrigues, C.H.M. & Ascher, D.B. CSM-Potential: mapping protein interactions and biological ligands in 3D space using geometric deep learning. Nucleic Acids Res 50, W204–W209 (2022).

98. Ittisoponpisan, S. et al. Can Predicted Protein 3D Structures Provide Reliable Insights into whether Missense Variants Are Disease Associated? J Mol Biol 431, 2197–2212 (2019).

99. Quinodoz, M. et al. AutoMap is a high performance homozygosity mapping tool using next-generation sequencing data. Nat Commun 12, 518 (2021).

100. Ware, J.S., Samocha, K.E., Homsy, J. & Daly, M.J. Interpreting de novo Variation in Human Disease Using denovolyzeR. Curr Protoc Hum Genet 87, 7 25 1–7 25 15 (2015).

101. Homsy, J. et al. De novo mutations in congenital heart disease with neurodevelopmental and other congenital anomalies. Science 350, 1262–6 (2015).

102. Doncheva, N.T., Morris, J.H., Gorodkin, J. & Jensen, L.J. Cytoscape StringApp: Network Analysis and Visualization of Proteomics Data. J Proteome Res 18, 623–632 (2019).

103. Shannon, P. et al. Cytoscape: a software environment for integrated models of biomolecular interaction networks. Genome Res 13, 2498–504 (2003).

104. Morris, J.H. et al. clusterMaker: a multi-algorithm clustering plugin for Cytoscape. BMC Bioinformatics 12, 436 (2011).

105. Huang da, W., Sherman, B.T. & Lempicki, R.A. Bioinformatics enrichment tools: paths toward the comprehensive functional analysis of large gene lists. Nucleic Acids Res 37, 1–13 (2009).

106. Huang da, W., Sherman, B.T. & Lempicki, R.A. Systematic and integrative analysis of large gene lists using DAVID bioinformatics resources. Nat Protoc 4, 44–57 (2009).

107. Chen, J., Bardes, E.E., Aronow, B.J. & Jegga, A.G. ToppGene Suite for gene list enrichment analysis and candidate gene prioritization. Nucleic Acids Res 37, W305–11 (2009).

108. Itan, Y. et al. The human gene connectome as a map of short cuts for morbid allele discovery. Proc Natl Acad Sci U S A 110, 5558–63 (2013).

109. Paradis, E., Claude, J. & Strimmer, K. APE: Analyses of Phylogenetics and Evolution in R language. Bioinformatics 20, 289–90 (2004).

110. Huang, D.W. et al. The DAVID Gene Functional Classification Tool: a novel biological module-centric algorithm to functionally analyze large gene lists. Genome Biol 8, R183 (2007).

111. Supek, F., Bosnjak, M., Skunca, N. & Smuc, T. REVIGO summarizes and visualizes long lists of gene ontology terms. PLoS One 6, e21800 (2011).

112. Langfelder, P. & Horvath, S. WGCNA: an R package for weighted correlation network analysis. BMC Bioinformatics 9, 559 (2008).

113. Walker, R.L. et al. Genetic Control of Expression and Splicing in Developing Human Brain Informs Disease Mechanisms. Cell 179, 750–771 e22 (2019).

114. Jin, S.C. et al. Exome sequencing implicates genetic disruption of prenatal neuro-gliogenesis in sporadic congenital hydrocephalus. Nat Med 26, 1754–1765 (2020).

115. Yu, G., Wang, L.G., Han, Y. & He, Q.Y. clusterProfiler: an R package for comparing biological themes among gene clusters. OMICS 16, 284–7 (2012).

116. Polioudakis, D. et al. A Single-Cell Transcriptomic Atlas of Human Neocortical Development during Mid-gestation. Neuron 103, 785–801 e8 (2019).

117. Lewis, S.A. et al. Mutation in ZDHHC15 Leads to Hypotonic Cerebral Palsy, Autism, Epilepsy, and Intellectual Disability. Neurol Genet 7, e602 (2021).

118. Hu, Y. et al. An integrative approach to ortholog prediction for disease-focused and other functional studies. BMC Bioinformatics 12, 357 (2011).

119. Mumberg, D., Muller, R. & Funk, M. Yeast vectors for the controlled expression of heterologous proteins in different genetic backgrounds. Gene 156, 119–22 (1995).

120. Ito, H., Fukuda, Y., Murata, K. & Kimura, A. Transformation of intact yeast cells treated with alkali cations. J Bacteriol 153, 163–8 (1983).

121. Harris, P.A. et al. Research electronic data capture (REDCap)--a metadata-driven methodology and workflow process for providing translational research informatics support. J Biomed Inform 42, 377–81 (2009).

122. Harris, P.A. et al. The REDCap consortium: Building an international community of software platform partners. J Biomed Inform 95, 103208 (2019).

123. Friedman, J.M. Neurofibromatosis 1. in GeneReviews((R)) (eds. Adam, M.P. et al.) (Seattle (WA), 1993).

124. Minchole, A. & Rodriguez, B. Artificial intelligence for the electrocardiogram. Nat Med 25, 22–23 (2019).

