## Supplementary Figures for "Recessive genomic and phenotypic variation in consanguineous families with cerebral palsy"

**Supplementary Figure 1 – QQ plot of observed vs. expected p-values showing enrichment of *de novo* muation in genes**
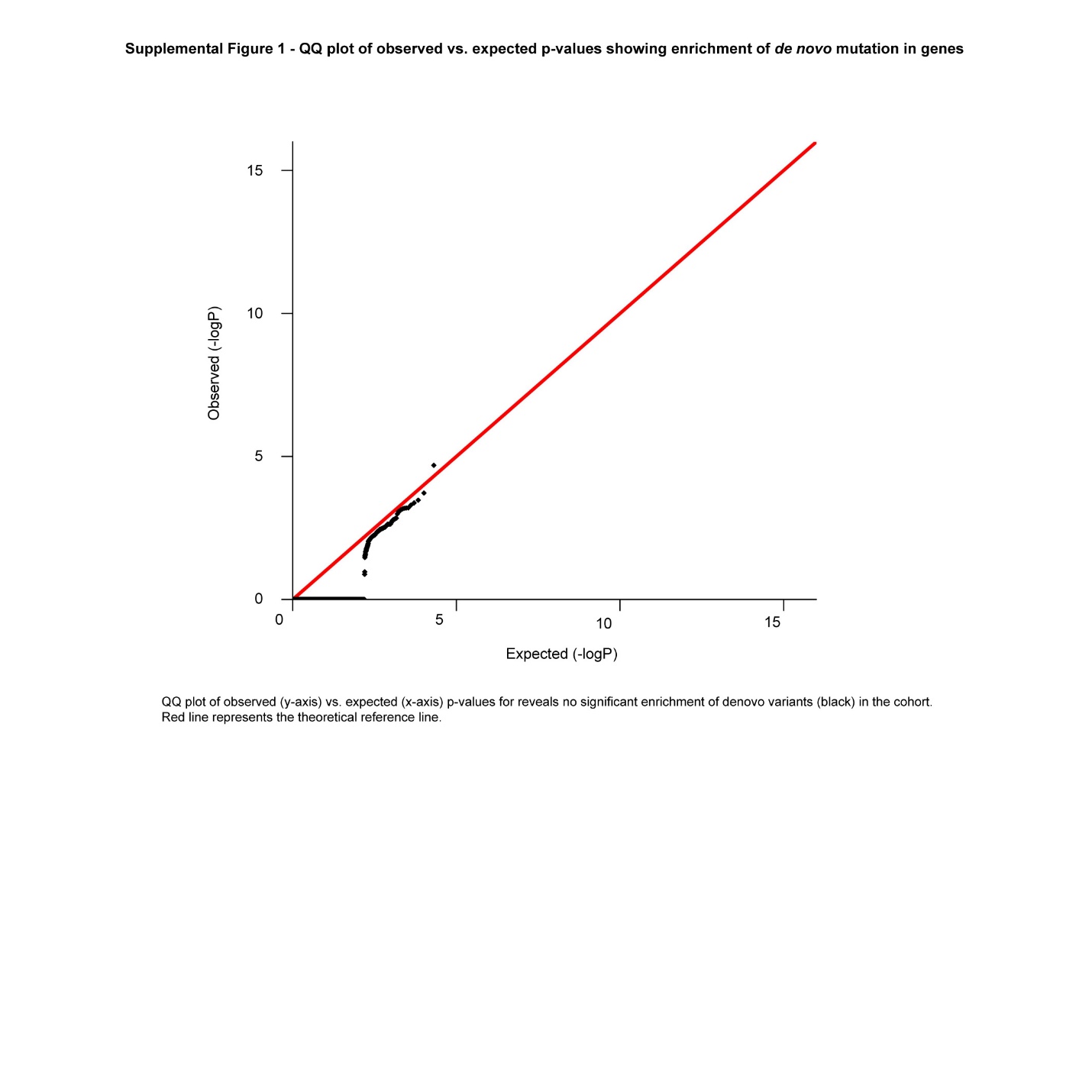

QQ plot of observed (y-axis) vs. expected p-values from *de novo* enrichment analysis reveals no significant enrichment of *de novo* variants (black points) in the cohort. Red line represents the theoretical reference line

**Supplementary Figure 2 – Comparisons between wild-type and mutant protein structures using normal mode analysis (DynaMut2) TCP1-MPND**
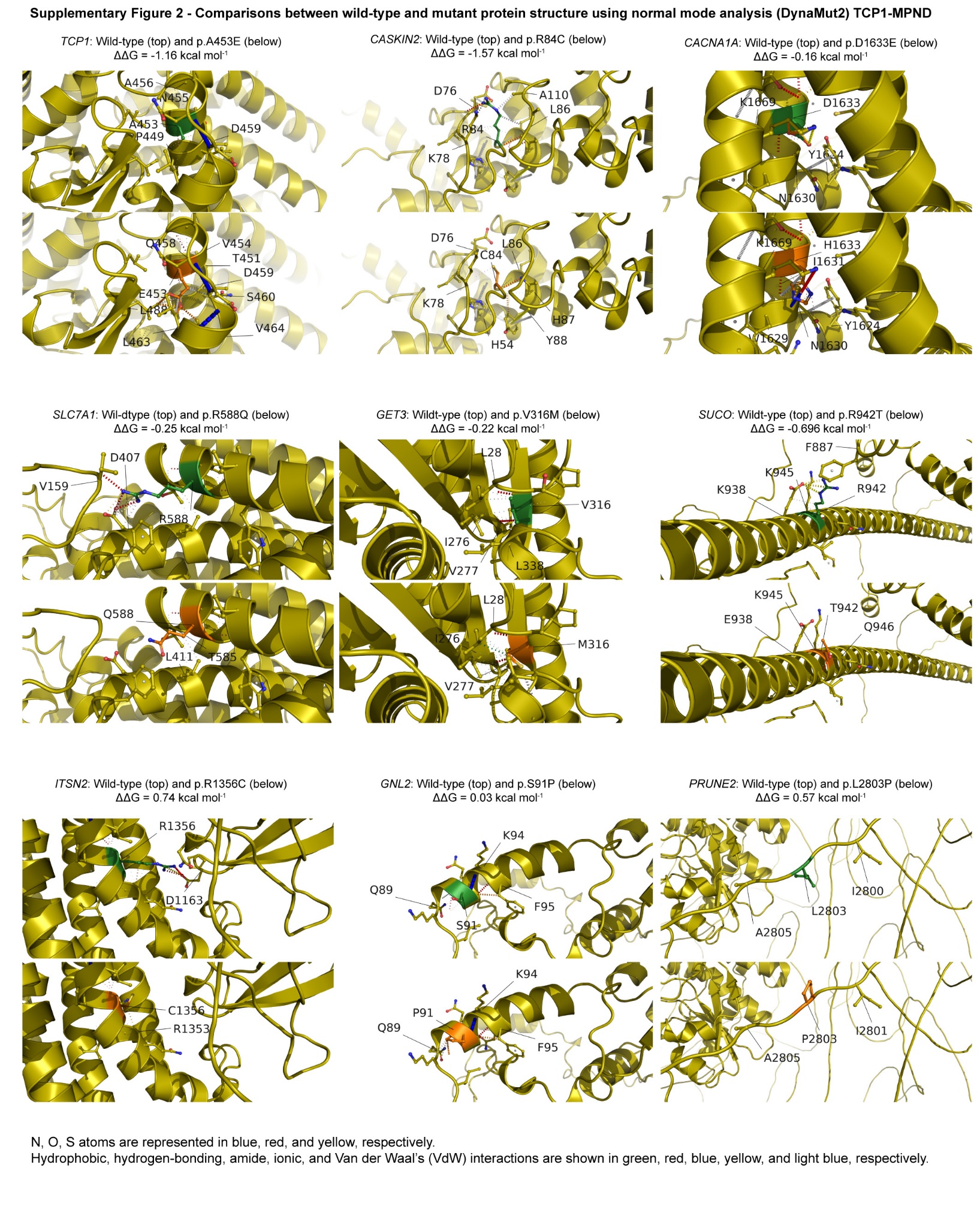

N, O, S atoms are represented in blue, red, and yellow, respectively. Hydrophobic, hydrogen-bonding, amide, ionic, and Van der Waal’s (VdW) interactions are shown in green, red, blue, yellow, and light blue, respectively. The wild-type residue is indicated in green, and the substituted residue is shown in orange.

**Supplementary Figure 2 – Comparisons between wild-type and mutant protein structures using normal mode analysis (DynaMut2) NDC1-WWC1**
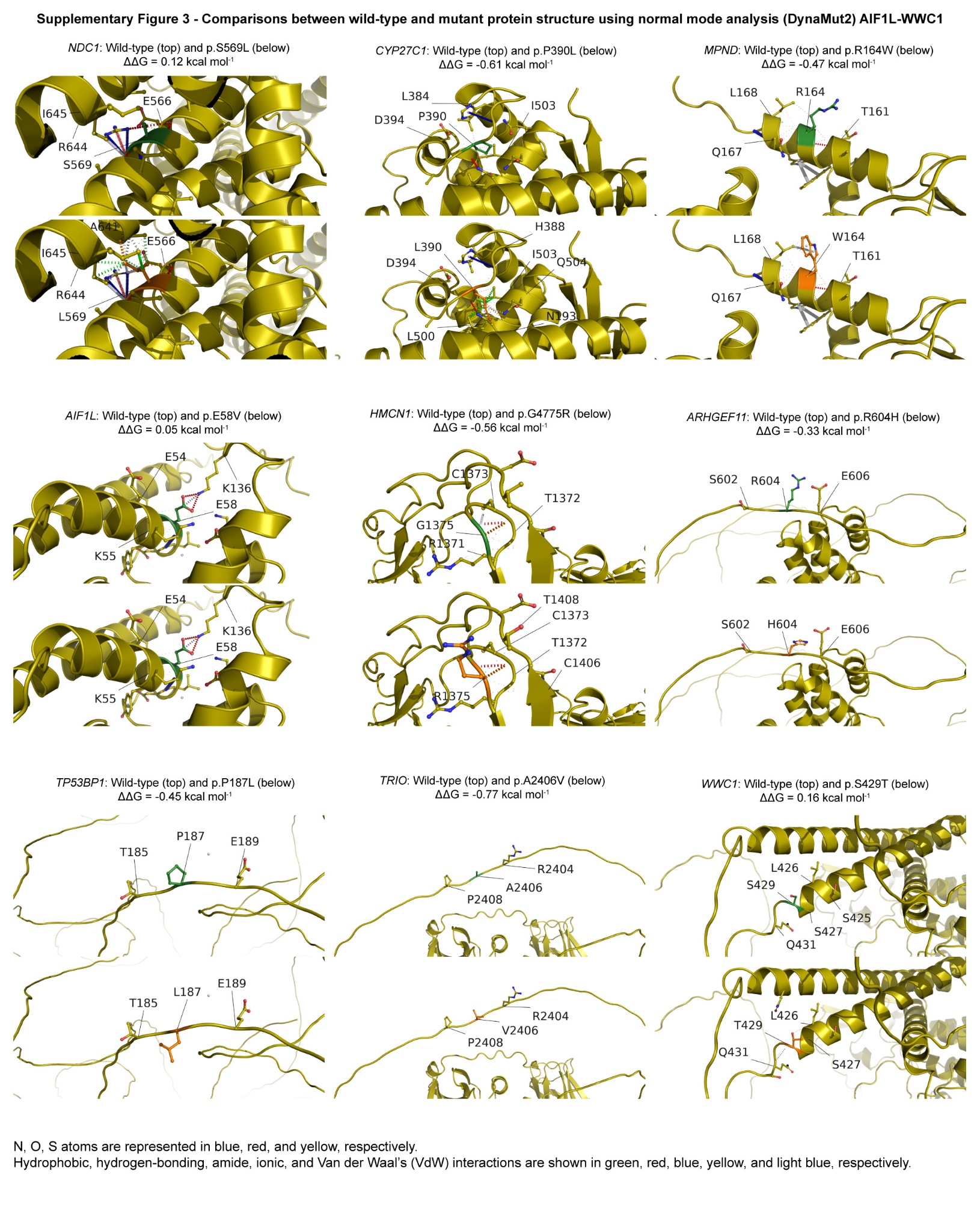

N, O, S atoms are represented in blue, red, and yellow, respectively. Hydrophobic, hydrogen-bonding, amide, ionic, and Van der Waal’s (VdW) interactions are shown in green, red, blue, yellow, and light blue, respectively. The wild-type residue is indicated in green, and the substituted residue is shown in orange.

**Supplementary Figure 4 – Weighted gene Co-expression network analysis (WGCNA) module enrichment**
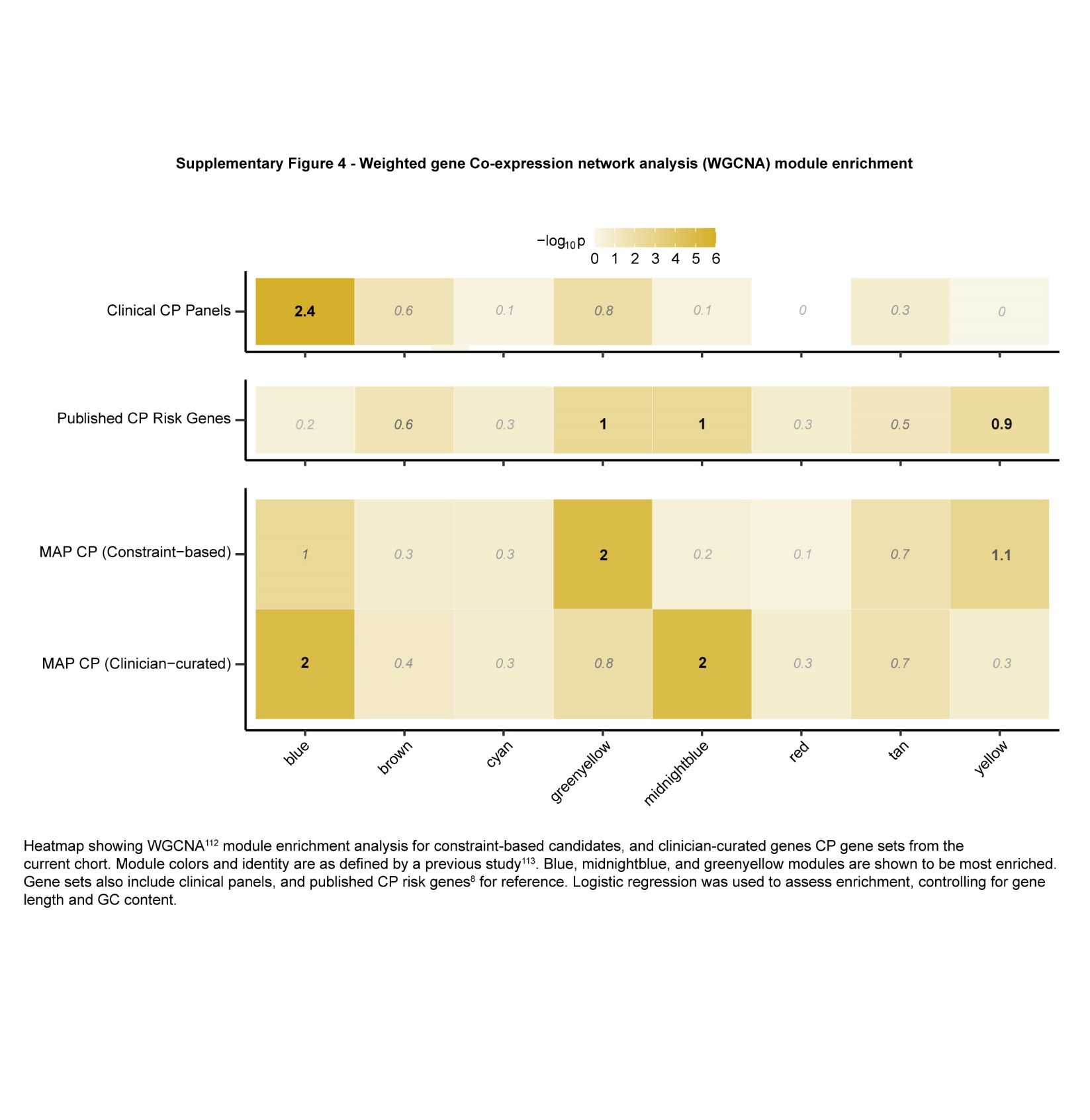
 Heatmap showing WGCNA^112^ module enrichment analysis for constraint-based candidates, and clinician-curated genes CP gene sets from the current cohort. Module colors and identity are as defined by a previous study.^113^ Blue, midnightblue, and greenyellow modules are shown to be most enriched.

Gene sets also include clinical panels, and published CP risk genes^8^ for reference. Logistic regression was used to assess enrichment, controlling for gene length and GC content.

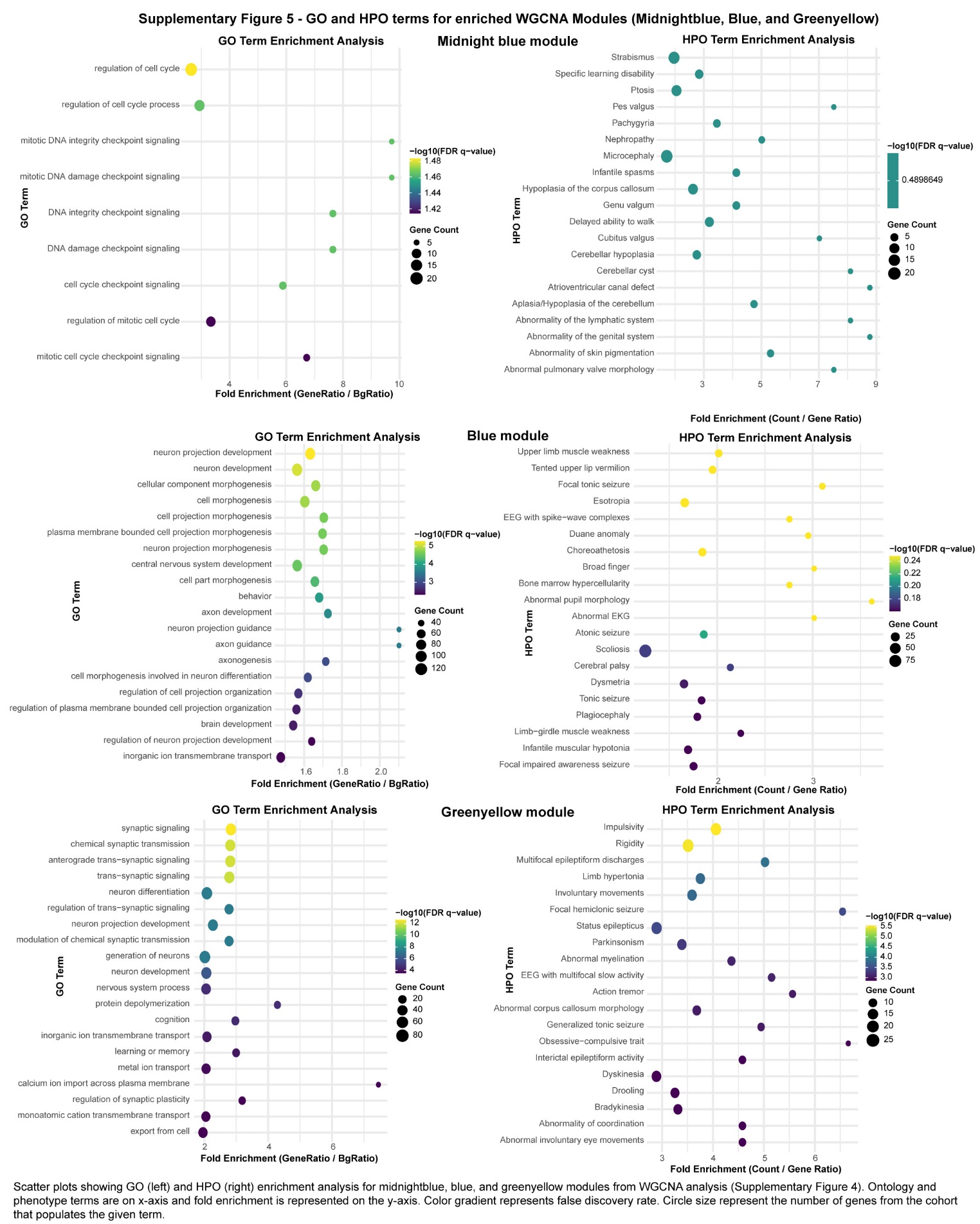
**Supplementary Figure 5 – GO and HPO terms for enriched WGCNA Modules (Midnightblue, Blue, and Greenyellow)**

Scatter plots showing GO (left) and HPO (right) enrichment analysis for midnightblue, blue, and greenyellow modules from WGCNA analysis (Supplementary Figure 4). Ontology and phenotype terms are on x-axis and fold enrichment is represented on the y-axis. Color gradient represents false discovery rate (FDR). Circle size represent the number of genes from the cohort that populates the given term.

**Supplementary Figure 6 – Distribution of comorbidities within the cohort**

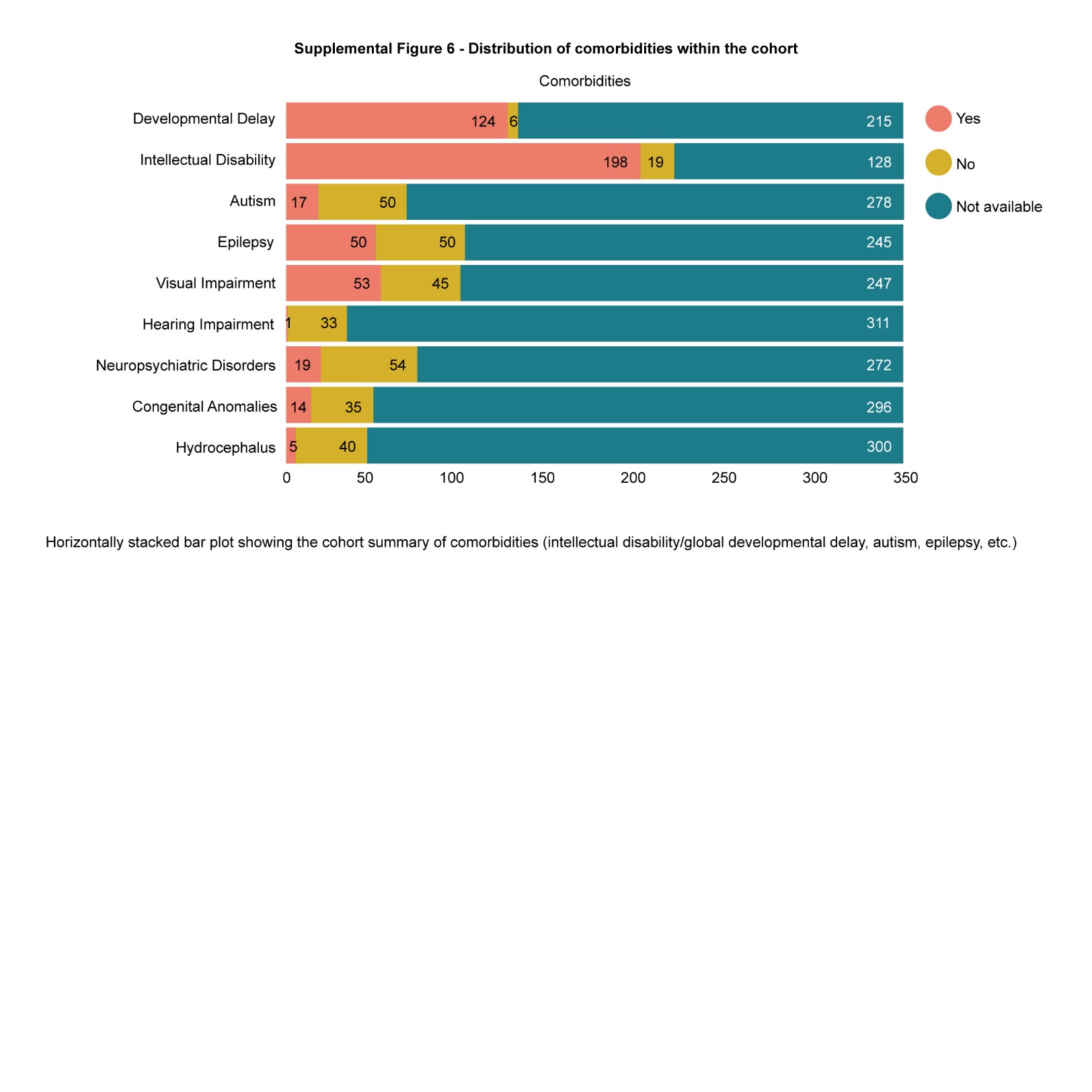
Horizontally stacked barplot showing the cohort summary of comorbidities (intellectual disability/global developmental delay, autism, epilepsy, etc.)
